# Tremor Improvement Despite Heterogeneous Ventral Intermediate Nucleus Targeting in Deep Brain Stimulation: A Systematic Review and Meta-Analysis

**DOI:** 10.64898/2026.04.07.26350347

**Authors:** Farzan Fahim, MohammadAmin Farajzadeh, Deniz Pourkhalil, Sadra Abedinzadeh, Reihaneh Ghahremani, Amirmahdi Mojtahedzadeh, Mojtaba Esmaeeli, Taha Mahdian, Dina Seyedi, Fatemeh Salarifar, Salar Pirbabaee, Sana Arbabi, Alireza Sedghi, Sayeh Oveisi, Guive Sharifi, Alireza Zali

**Author notes:** Corresponding author: Farzan Fahim.

## Abstract

**Background:** Deep brain stimulation (DBS) targeting the ventral intermediate nucleus (Vim) of the thalamus is an established surgical therapy for medically refractory tremor, particularly essential tremor. Accurate localization of the Vim remains challenging because the nucleus is not directly visible on conventional MRI. Consequently, multiple targeting approaches have been developed, including atlas-based stereotactic coordinates, microelectrode recording (MER), advanced MRI visualization techniques, and diffusion-based tractography. This systematic review and meta-analysis evaluated current Vim targeting strategies and synthesized tremor outcomes following intervention.

**Methods:** This systematic review and meta-analysis was conducted according to PRISMA 2020 guidelines and registered in PROSPERO. PubMed/MEDLINE, Scopus, Web of Science, and Embase were searched from inception to January 29, 2026. Studies investigating Vim-targeted tremor surgery and reporting targeting strategies or tremor outcomes were eligible. Data extraction and risk-of-bias assessment were performed independently by two reviewers using JBI and QUADAS-2 tools. Random-effects meta-analysis using standardized mean differences (Hedges’ g) was performed to evaluate pre- to postoperative tremor improvement.

**Results:** A total of 2,398 records were identified, and 25 studies met inclusion criteria for the systematic review. Across these studies, 211 patients undergoing Vim-targeted tremor surgery were analyzed. Considerable heterogeneity was observed in study design, patient populations, imaging protocols, and targeting approaches, including atlas-based targeting, MER-guided localization, advanced MRI visualization, and diffusion tractography of tremor-related pathways such as the dentato-rubro-thalamic tract. Six studies comprising seven independent cohorts provided sufficient data for meta-analysis. Pooled analysis demonstrated substantial tremor improvement following intervention (SMD −3.91, 95% CI −4.81 to −3.01; p < 0.0001). Although between-study heterogeneity was moderate to substantial (Q = 18.12, p = 0.0059; I² = 66.9%), all cohorts showed consistent reductions in tremor severity. Sensitivity analyses confirmed the stability of the pooled effect, and funnel plot and trim-and-fill analyses did not indicate significant publication bias.

**Conclusions:** Despite substantial heterogeneity in Vim targeting methodologies, surgical intervention consistently produces marked tremor reduction. Across anatomical, electrophysiological, and imaging-based targeting approaches, clinical outcomes remained robust. Future prospective studies with standardized outcome reporting and direct comparisons of targeting techniques are needed to determine whether emerging imaging-guided strategies provide measurable clinical advantages.

## Introduction

Deep brain stimulation (DBS) has emerged as an effective neuromodulatory treatment for a range of functional neurological disorders, particularly movement disorders characterized by disabling tremor. Tremor is a common clinical manifestation in several neurological conditions, including essential tremor (ET), Parkinson’s disease (PD), and other disorders such as multiple sclerosis. In patients with medically refractory tremor, DBS has demonstrated sustained efficacy and has become an established therapeutic option in clinical practice(1, 2). Several neural targets have been explored for tremor control depending on the underlying disease. For example, stimulation of the subthalamic nucleus (STN) and globus pallidus internus (GPi) has shown effectiveness for tremor in Parkinson’s disease, while stimulation of the fornix has been investigated in Alzheimer’s disease with modest motor benefits(3, 4). Nevertheless, the ventral intermediate (VIM) nucleus of the thalamus remains the principal and most widely used surgical target for tremor control, particularly in essential tremor(5).

Accurate localization of the VIM nucleus is therefore a critical step in DBS procedures. However, the VIM nucleus cannot be reliably visualized on conventional structural magnetic resonance imaging, which has led to the development of several targeting strategies. Traditionally, indirect targeting methods based on stereotactic anatomical landmarks identified on computed tomography (CT) or MRI have been used to estimate the location of the VIM(2, 6). More recently, connectivity-based approaches such as probabilistic thalamic segmentation have been introduced, enabling identification of thalamic nuclei according to their cortical connectivity patterns(6).

Advances in neuroimaging have enabled more direct targeting strategies for the VIM. Functional MRI at 3 T has been used to localize the dentatorubrothalamic tract (DRT), a major cerebellothalamic pathway terminating within the VIM(7). Diffusion MRI tractography has further facilitated visualization of tremor-related white matter pathways, leading to the development of tractography-guided targeting strategies, including single-tract (DRT), dual-tract (pyramidal and somatosensory), and multi-tract models(8). In addition, intraoperative microelectrode recordings may assist in refining target localization, and newer stereotactic systems such as the ClearPoint SmartFrame OR™ have demonstrated submillimeter targeting accuracy without requiring intraoperative MRI(9, 10).

Despite the development of multiple imaging-based and electrophysiological targeting techniques, there remains no clear consensus regarding which approach provides the most accurate localization of the VIM or yields superior clinical outcomes. Different methods require varying levels of technological resources, expertise, and cost, and current evidence has not definitively established the superiority of one technique over another.

Therefore, the aim of the present study was to conduct a comprehensive systematic review and meta-analysis of the existing literature evaluating different VIM targeting strategies in DBS for tremor. By synthesizing available evidence, this study seeks to determine which targeting approaches are most effective in reducing tremor severity and improving patient outcomes.

## METHODS

### Study Design and Registration

This systematic review and meta-analysis were conducted in accordance with the Preferred Reporting Items for Systematic Reviews and Meta-Analyses (PRISMA) 2020 guidelines(11). The PRISMA checklist is provided in Supplementary file 1. The study protocol was prospectively registered in the International Prospective Register of Systematic Reviews (PROSPERO)(12) under registration number CRD420261362914, which is provided in supplementory file 2.

### Search Strategy

A comprehensive systematic search was conducted across four major electronic databases including PubMed, Scopus, Web of Science, and Embase. The search covered the entire time span of each database from inception to January 29, 2026. No language or publication date restrictions were applied. For studies published in languages other than English, translation was planned using reliable online translation tools to allow accurate evaluation during the screening process.

The complete search syntax for all databases is provided in Supplementary file 3. The full PubMed search strategy used in this review was as follows:

((“ventral intermediate nucleus”[tiab] OR VIM[tiab] OR “ventralis intermedius”[tiab] OR “thalamic nucleus”[tiab])

AND

(segmentation[tiab] OR parcellation[tiab] OR targeting[tiab] OR localization[tiab] OR atlas[tiab] OR tractography[tiab] OR “diffusion MRI”[tiab] OR connectivity[tiab] OR “automated segmentation”[tiab] OR “artificial intelligence”[tiab] OR “electrophysiologic mapping”[tiab] OR “brain mapping”[tiab] OR mapping[tiab] OR parcel*[tiab] OR segment*[tiab] OR tractograph*[tiab] OR diffus*[tiab] OR connect*[tiab] OR fMRI[tiab] OR FGATIR[tiab] OR “fast gray matter acquisition T1 inversion recovery”[tiab])

AND

(“deep brain stimulation”[tiab] OR DBS[tiab])

### Eligibility Criteria

Study eligibility was defined according to predefined inclusion and exclusion criteria. Studies were eligible for inclusion if they investigated patients undergoing deep brain stimulation (DBS) with targeting of the ventral intermediate nucleus (Vim) of the thalamus. The exposure of interest was Vim targeting, while the comparator consisted of different anatomical, imaging-based, or physiological targeting strategies used to localize the Vim. The primary outcome of interest was the comparative effectiveness or accuracy of different targeting strategies, particularly in relation to tremor outcomes or targeting accuracy. Eligible study designs included cross-sectional studies, cohort studies (prospective or retrospective), case-control studies, and case series. Studies were excluded if they were case reports, narrative or systematic reviews, conference abstracts, or technical notes. Case series including fewer than 10 patients were also excluded. In addition, studies that did not specifically evaluate Vim targeting strategies or did not involve DBS procedures were excluded.

### Study Selection

All retrieved records were imported into EndNote version 21 for reference management and duplicate removal. Title and abstract screening was performed independently by two reviewers. A comprehensive Excel spreadsheet documenting excluded studies was prepared by the senior author (FF). This file included the article title, first author, DOI, and the reason for exclusion and is provided as Supplementary file 4.

Following the title and abstract screening phase, full-text assessment was conducted. Because 140 studies progressed to the full-text screening stage, a pilot calibration step was implemented to assess reviewer agreement. Initially, 28 studies were randomly selected by MAF and independently screened by two reviewers (SA and DP). Inter-reviewer agreement was calculated and showed a concordance rate of 73%, which was below the predefined threshold of 80% set by the senior author. Consequently, the remaining studies were screened independently by the same reviewers.

During the screening process, each reviewer independently completed two structured Excel forms prepared by the senior author. One form contained information for included studies, including title, first author, publication year, study design, target population, Vim targeting method, and study outcomes. The second form documented excluded studies and included article title, author, country of origin, and the reason for exclusion. These files are provided in Supplementary file 5 and 6. Disagreements between reviewers were resolved through discussion and final adjudication by the senior author (FF).

### Data Extraction

A standardized data extraction form was developed by the senior author (FF) prior to data collection. Following training on the extraction procedure, two reviewers (DP and ME) independently extracted data from each eligible study. The extracted data were subsequently verified and finalized by the senior author. The data extraction sheet included detailed study-level, patient-level, imaging, surgical, and outcome variables. These included bibliographic information (title, authors, publication year, DOI, funding source), study characteristics (country, center type, study design, data collection period), eligibility criteria, follow-up duration, and ethical approval status. Patient-level variables included sample size, mean age, gender distribution, disease type, and disease duration. Imaging and targeting variables included the primary method used for Vim border determination, MRI field strength, MRI sequences used, voxel resolution, direct visualization methods, diffusion tractography approaches, software pipelines, regions of interest, and pathway definitions used for Vim localization. Intraoperative variables included the number of microelectrode recording (MER) trajectories, depth of recordings, physiological criteria used to identify the Vim, sensory responses, and microstimulation findings. Atlas-based targeting variables included atlas name, atlas version, reference space, registration methods, software used for image registration, probability thresholds, and registration errors. Surgical accuracy variables included target coordinates, AC-PC measurements, distance from midline, and lead placement accuracy metrics such as Euclidean error, radial error, and depth error. Clinical outcome variables included preoperative and postoperative tremor scores, percent improvement, stimulation thresholds required for tremor control, and adverse events related to electrode misplacement. All extracted datasets are provided in Supplementary file 7.

### Risk of Bias Assessment

Given the methodological diversity of the included studies, two different risk-of-bias assessment tools were employed. Observational cohort studies were evaluated using the Joanna Briggs Institute (JBI) Critical Appraisal Checklist for Cohort Studies. Imaging methodological studies were assessed using the QUADAS-2 (Quality Assessment of Diagnostic Accuracy Studies) tool. Following training on the assessment procedures conducted by MAF, two reviewers (SP and FS) independently performed the risk-of-bias evaluation for each included study. Any disagreements were resolved through discussion and consensus. The full risk-of-bias assessment results are presented in Supplementary file 8.

### Data Synthesis and Statistical Analysis

Quantitative synthesis was performed using R statistical software (version 4.5.1; R Foundation for Statistical Computing, Vienna, Austria). Meta-analysis procedures were conducted using established evidence-synthesis libraries, including the metafor, meta, and dmetar packages, which provide validated implementations for random-effects modeling, influence diagnostics, and publication bias assessment. Eligibility for meta-analysis was determined after evaluation of the extracted dataset. Because tremor outcomes were reported using different clinical rating scales across the included studies, most commonly the Fahn–Tolosa–Marin Tremor Rating Scale (FTMTRS) and The Essential Tremor Rating Assessment Scale (TETRAS), effect sizes were standardized to allow cross-study comparison. Accordingly, pooled estimates were calculated using the standardized mean difference (SMD) with corresponding 95% confidence intervals (CI). Effect sizes were calculated using the Hedges’ g formulation, which corrects for small-sample bias and is recommended for meta-analyses involving relatively small study populations. For each study, the standardized mean difference was calculated as:

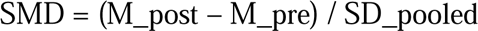

where M_post represents the postoperative tremor score and M_pre represents the preoperative tremor score.

The pooled standard deviation was calculated as:

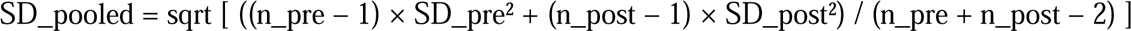

To reduce small-sample bias, the effect size was multiplied by the Hedges correction factor:

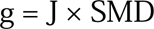

where

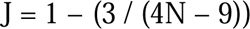

and N represents the total sample size.

The variance of the standardized mean difference was estimated as:

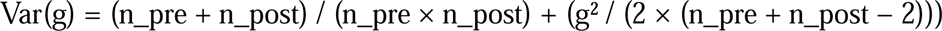

These calculations were implemented through the metafor package using the escalc() function, ensuring consistent estimation of effect sizes and variances across studies. Because outcome values of zero were observed in some extracted datasets, a continuity correction of 0.5 was applied where necessary to allow computation of standardized effects and avoid undefined variance estimates. Given the substantial methodological heterogeneity identified among included studies, including differences in imaging approaches, targeting strategies, patient populations, and outcome scales, a random-effects model was used for pooled analysis. Random-effects models were estimated using the restricted maximum likelihood (REML) estimator implemented in the metafor package. Between-study heterogeneity was quantified using both Cochran’s Q statistic and the I² statistic, where I² values of approximately 25%, 50%, and 75% were interpreted as representing low, moderate, and high heterogeneity, respectively. Forest plots were generated using the meta and metafor packages to visually display individual study effect sizes and pooled estimates with corresponding confidence intervals. To evaluate the robustness of the pooled estimates, several sensitivity and diagnostic analyses were conducted. A leave-one-out sensitivity analysis was performed using the leave1out() function in the metafor package to determine whether the pooled effect size was disproportionately influenced by any single study. In addition, influence diagnostics were conducted using the influence () function to identify potentially influential observations based on Cook’s distance, DFBETAS, and covariance ratios. Potential publication bias was explored through visual inspection of funnel plots and further evaluated using trim-and-fill analysis implemented via the trimfill() function.

## RESULTS

### A full version of Result is provided in supplementory file 9

#### Study Selection

The study selection process is illustrated in the PRISMA flow diagram (Figure 1). A total of 2,398 records were identified through database searching. After removal of 1,016 duplicate records, 1,382 studies remained for title and abstract screening. During this stage, 1,235 records were excluded for the following reasons: conference abstracts, protocols, or registry-only reports (n = 70), studies not relevant to the predefined PICOS criteria (n = 570), and studies with inappropriate design or population (n = 595). A total of 147 full-text reports were sought for retrieval, of which seven could not be obtained. Consequently, 140 full-text articles were assessed for eligibility. Among these, several studies were excluded for the following reasons: case series with fewer than 10 participants (n = 11), studies targeting nuclei other than the ventral intermediate nucleus (Vim) (n = 50), studies employing irrelevant methodological approaches (n = 28), studies not specifically addressing Vim targeting (n = 20), and studies with ineligible designs (n = 6).Ultimately, 25 studies met the eligibility criteria and were included in the systematic review. These comprised 14 retrospective cohort studies, 3 prospective cohort studies, 1 cross-sectional observational study, and 7 methodological or imaging studies focusing on Vim localization. Among the included studies, 6 studies (7 cohorts) provided sufficient quantitative data on preoperative and postoperative tremor scores and were therefore included in the quantitative meta-analysis.

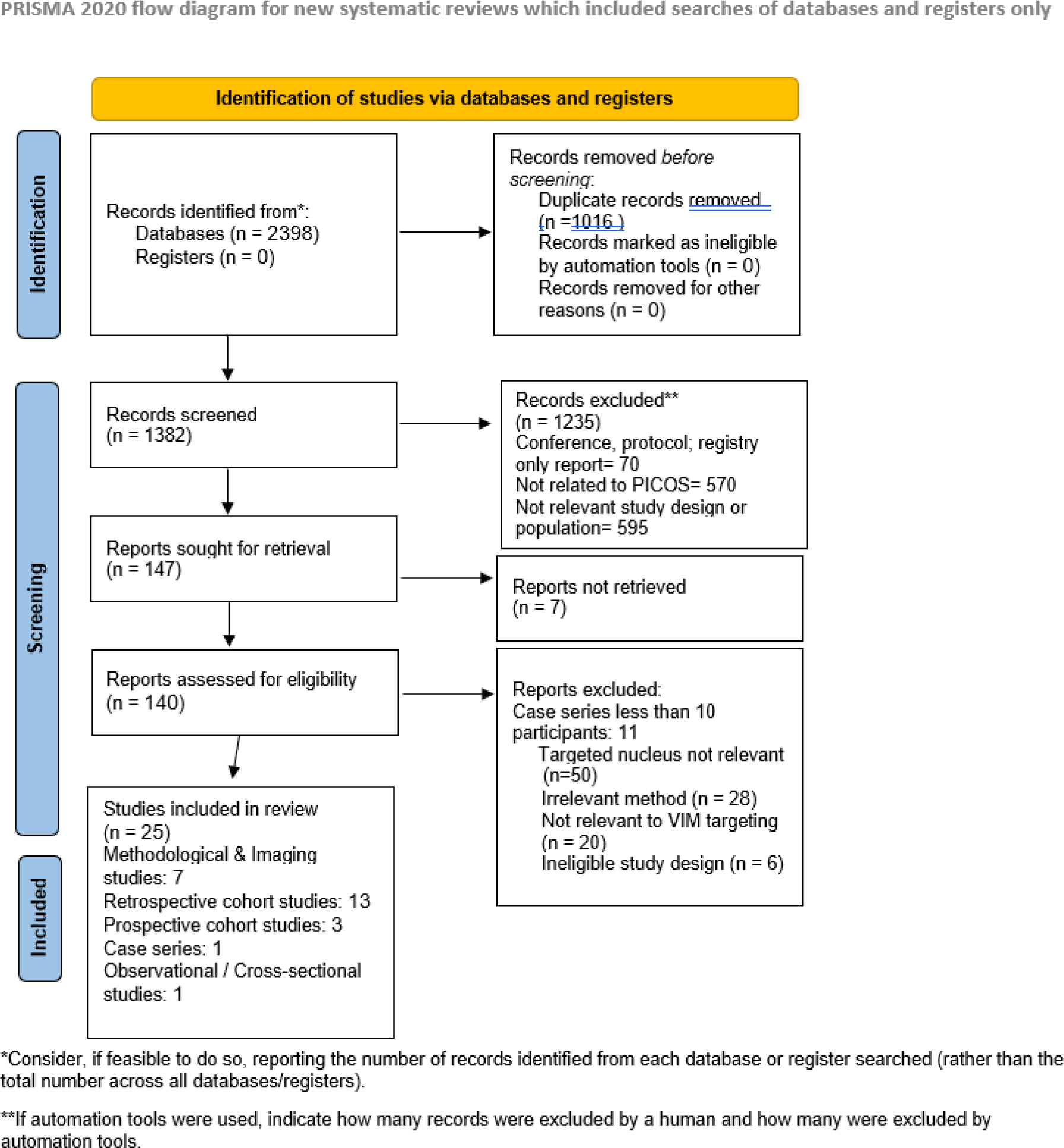

### Study Characteristics

The included studies demonstrated substantial heterogeneity in study design, patient populations, imaging protocols, and approaches used for localization of the ventral intermediate nucleus of the thalamus. Study designs ranged from retrospective observational cohorts and prospective clinical studies to imaging methodological investigations evaluating novel visualization or segmentation techniques. Several studies evaluated clinical outcomes of deep brain stimulation (DBS) targeting the Vim. For example, Vassal et al(13). reported a retrospective case series of patients undergoing Vim DBS for medically refractory tremor, while Burchiel et al(14). conducted a prospective cohort study assessing stereotactic accuracy of image-guided DBS implantation using intraoperative computed tomography. Other investigations used comparative designs to evaluate different targeting strategies. Fenoy et al 2018 (15) compared tractography-guided targeting of the dentato-rubro-thalamic tract (DRTT) with conventional atlas-based Vim targeting in patients with essential tremor. Patient populations varied across studies but were largely composed of individuals with medically refractory tremor syndromes. Essential tremor represented the most common diagnosis, although several cohorts also included patients with Parkinson disease tremor or mixed tremor etiologies. In addition, a subset of studies consisted of imaging methodological investigations performed in healthy participants or mixed populations in order to evaluate thalamic segmentation algorithms or connectivity-based localization techniques. Substantial variability was also observed in imaging approaches. Conventional stereotactic targeting using anterior commissure–posterior commissure (AC–PC) coordinates and stereotactic atlases remained widely used. However, several studies investigated advanced imaging techniques designed to improve visualization of thalamic nuclei. Examples include white-matter-attenuated inversion recovery (WAIR) imaging, ultra-high-field 7-Tesla MRI, diffusion tensor imaging (DTI), and diffusion-based connectivity mapping. Outcome reporting also differed between studies. Clinical outcomes were assessed using tremor severity scales such as the Fahn–Tolosa–Marin Tremor Rating Scale (FTMTRS) or functional outcome measures including activities of daily living scores. Follow-up durations ranged from short-term postoperative assessments of several months to long-term follow-up exceeding three years. A summary of the included studies is provided in Table 1.

**Table 1.**
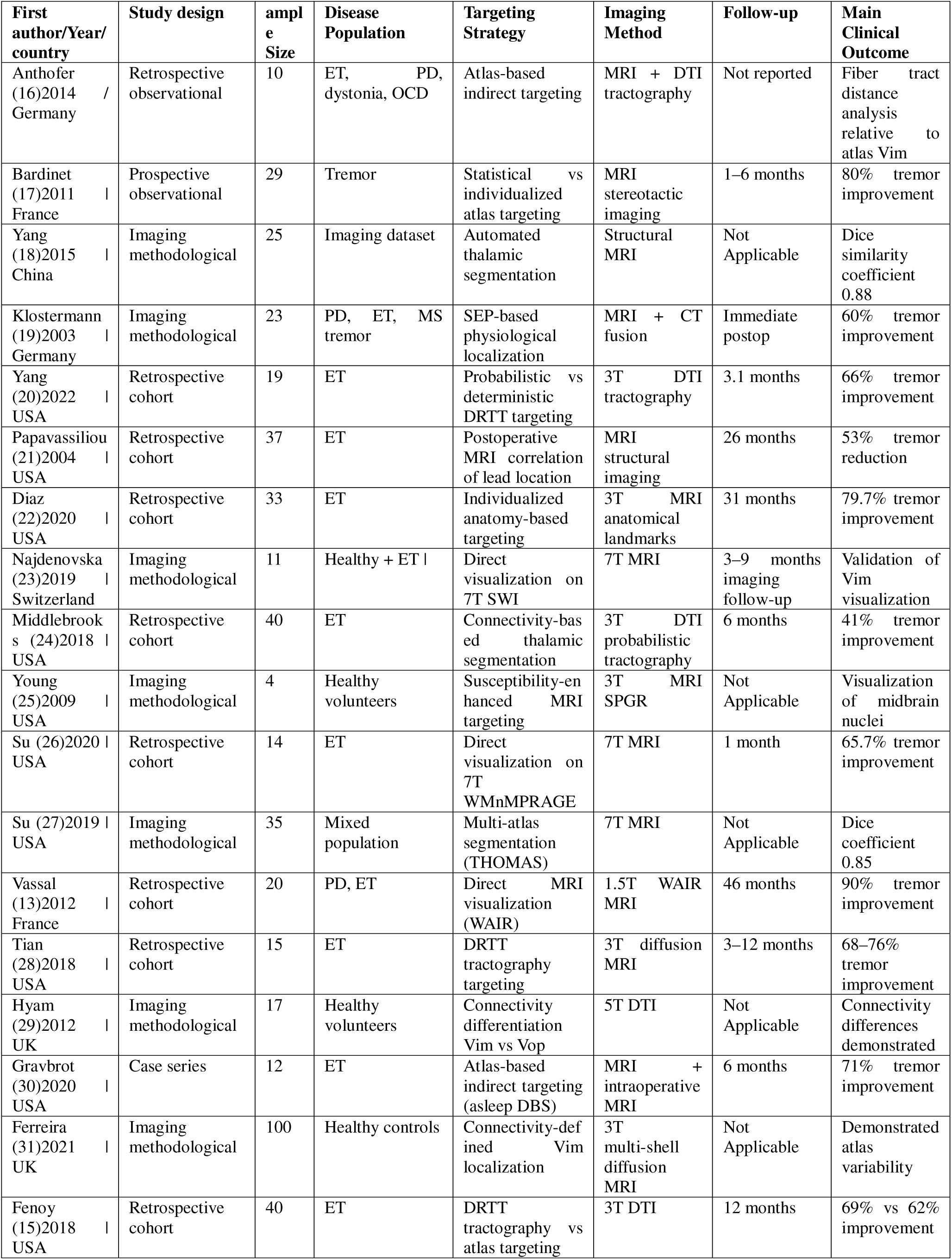
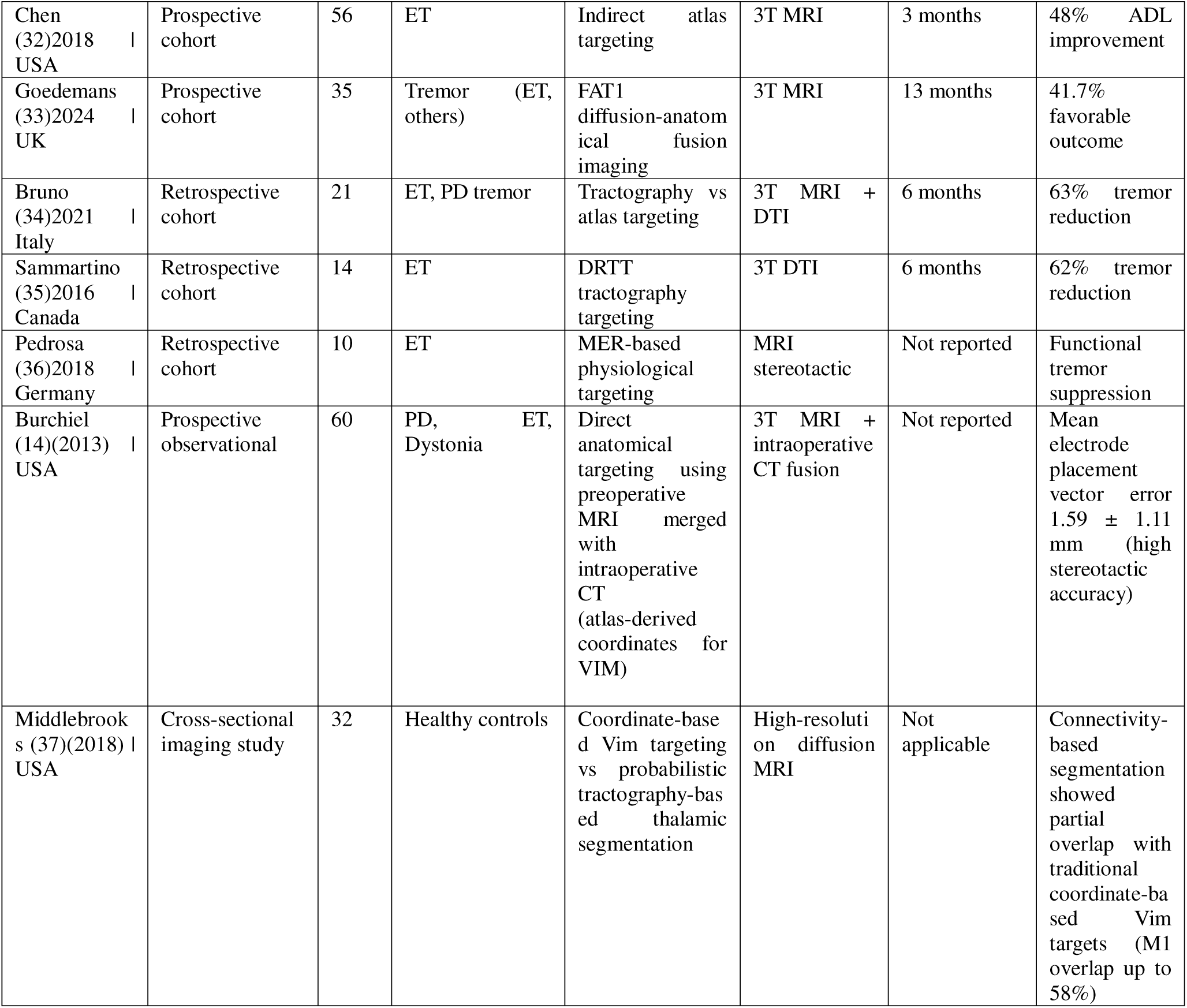
summary of included studies.

| First author/Year/<br>country | Study design | sample<br>Size | Disease<br>Population | Targeting<br>Strategy | Imaging<br>Method | Follow-up | Main<br>Clinical<br>Outcome |
| --- | --- | --- | --- | --- | --- | --- | --- |
| Anthofer<br>(16)2014 /<br>Germany | Retrospective<br>observational | 10 | ET, PD,<br>dystonia, OCD | Atlas-based<br>indirect targeting | MRI + DTI<br>tractography | Not reported | Fiber tract<br>distance<br>analysis<br>relative to<br>atlas Vim |
| Bardinet<br>(17)2011 \|<br>France | Prospective<br>observational | 29 | Tremor | Statistical vs<br>individualized<br>atlas targeting | MRI<br>stereotactic<br>imaging | 1–6 months | 80% tremor<br>improvement |
| Yang<br>(18)2015 \|<br>China | Imaging<br>methodological | 25 | Imaging dataset | Automated<br>thalamic<br>segmentation | Structural<br>MRI | Not<br>Applicable | Dice<br>similarity<br>coefficient<br>0.88 |
| Klostermann<br>(19)2003 \|<br>Germany | Imaging<br>methodological | 23 | PD, ET, MS<br>tremor | SEP-based<br>physiological<br>localization | MRI + CT<br>fusion | Immediate<br>postop | 60% tremor<br>improvement |
| Yang<br>(20)2022 \|<br>USA | Retrospective<br>cohort | 19 | ET | Probabilistic vs<br>deterministic<br>DRTT targeting | 3T DTI<br>tractography | 3.1 months | 66% tremor<br>improvement |
| Papavassiliou<br>(21)2004 \|<br>USA | Retrospective<br>cohort | 37 | ET | Postoperative<br>MRI correlation<br>of lead location | MRI<br>structural<br>imaging | 26 months | 53% tremor<br>reduction |
| Diaz<br>(22)2020 \|<br>USA | Retrospective<br>cohort | 33 | ET | Individualized<br>anatomy-based<br>targeting | 3T MRI<br>anatomical<br>landmarks | 31 months | 79.7% tremor<br>improvement |
| Najdenovska<br>(23)2019 \|<br>Switzerland | Imaging<br>methodological | 11 | Healthy + ET \| | Direct<br>visualization on<br>7T SWI | 7T MRI | 3–9 months<br>imaging<br>follow-up | Validation of<br>Vim<br>visualization |
| Middlebrook<br>s (24)2018 \|<br>USA | Retrospective<br>cohort | 40 | ET | Connectivity-bas<br>ed thalamic<br>segmentation | 3T DTI<br>probabilistic<br>tractography | 6 months | 41% tremor<br>improvement |
| Young<br>(25)2009 \|<br>USA | Imaging<br>methodological | 4 | Healthy<br>volunteers | Susceptibility-en<br>hanced MRI<br>targeting | 3T MRI<br>SPGR | Not<br>Applicable | Visualization<br>of midbrain<br>nuclei |
| Su (26)2020 \|<br>USA | Retrospective<br>cohort | 14 | ET | Direct<br>visualization on<br>7T<br>WMnMPRAGE | 7T MRI | 1 month | 65.7% tremor<br>improvement |
| Su (27)2019 \|<br>USA | Imaging<br>methodological | 35 | Mixed<br>population | Multi-atlas<br>segmentation<br>(THOMAS) | 7T MRI | Not<br>Applicable | Dice<br>coefficient<br>0.85 |
| Vassal<br>(13)2012 \|<br>France | Retrospective<br>cohort | 20 | PD, ET | Direct MRI<br>visualization<br>(WAIR) | 1.5T WAIR<br>MRI | 46 months | 90% tremor<br>improvement |
| Tian<br>(28)2018 \|<br>USA | Retrospective<br>cohort | 15 | ET | DRTT<br>tractography<br>targeting | 3T diffusion<br>MRI | 3–12 months | 68–76%<br>tremor<br>improvement |
| Hyam<br>(29)2012 \|<br>UK | Imaging<br>methodological | 17 | Healthy<br>volunteers | Connectivity<br>differentiation<br>Vim vs Vop | 5T DTI | Not<br>Applicable | Connectivity<br>differences<br>demonstrated |
| Gravbrot<br>(30)2020 \|<br>USA | Case series | 12 | ET | Atlas-based<br>indirect targeting<br>(asleep DBS) | MRI +<br>intraoperative<br>MRI | 6 months | 71% tremor<br>improvement |
| Ferreira<br>(31)2021 \|<br>UK | Imaging<br>methodological | 100 | Healthy controls | Connectivity-def<br>ined Vim<br>localization | 3T<br>multi-shell<br>diffusion<br>MRI | Not<br>Applicable | Demonstrated<br>atlas<br>variability |
| Fenoy<br>(15)2018 \|<br>USA | Retrospective<br>cohort | 40 | ET | DRTT<br>tractography vs<br>atlas targeting | 3T DTI | 12 months | 69% vs 62%<br>improvement |
| Chen (32)2018 USA | Prospective cohort | 56 | ET | Indirect atlas targeting | 3T MRI | 3 months | 48% ADL improvement |
| Goedemans (33)2024 UK | Prospective cohort | 35 | Tremor (ET, others) | FAT1 diffusion-anatomical fusion imaging | 3T MRI | 13 months | 41.7% favorable outcome |
| Bruno (34)2021 Italy | Retrospective cohort | 21 | ET, PD tremor | Tractography vs atlas targeting | 3T MRI + DTI | 6 months | 63% tremor reduction |
| Sammartino (35)2016 Canada | Retrospective cohort | 14 | ET | DRTT tractography targeting | 3T DTI | 6 months | 62% tremor reduction |
| Pedrosa (36)2018 Germany | Retrospective cohort | 10 | ET | MER-based physiological targeting | MRI stereotactic | Not reported | Functional tremor suppression |
| Burchiel (14)(2013) USA | Prospective observational | 60 | PD, ET, Dystonia | Direct anatomical targeting using preoperative MRI merged with intraoperative CT (atlas-derived coordinates for VIM) | 3T MRI + intraoperative CT fusion | Not reported | Mean electrode placement vector error $1.59 \pm 1.11$ mm (high stereotactic accuracy) |
| Middlebrooks (37)(2018) USA | Cross-sectional imaging study | 32 | Healthy controls | Coordinate-based Vim targeting vs probabilistic tractography-based thalamic segmentation | High-resolution diffusion MRI | Not applicable | Connectivity-based segmentation showed partial overlap with traditional coordinate-based Vim targets (M1 overlap up to 58%) |

### Patient Characteristics

Across the included studies, a total of 211 patients undergoing Vim-targeted tremor surgery were analyzed. Most cohorts consisted of individuals with disabling tremor refractory to pharmacological therapy. Essential tremor was the predominant underlying diagnosis across studies, although several cohorts also included patients with Parkinson disease tremor. For instance, Vassal (13)et al. reported a cohort of 20 patients undergoing Vim DBS, including 13 patients with parkinson disease tremor and 7 patients with essential tremor. Similarly, the prospective study by Burchiel (14)et al. included patients undergoing DBS for movement disorders, among whom 25 individuals received Vim targeting. Other investigations focused exclusively on essential tremor populations, such as the cohort described by Chen (32)et al., which included 56 patients undergoing Vim DBS implantation. Demographic characteristics were broadly comparable across studies, with most cohorts consisting of older adults. Reported mean age ranged from approximately 61 to 68 years. Male patients were generally more prevalent than female patients in the available cohorts. Surgical approaches varied between unilateral and bilateral implantation strategies. In the Vassal (13)et al. series, bilateral DBS implantation was performed in 12 patients and unilateral implantation in 8 patients. Similarly, other cohorts reported mixed unilateral and bilateral procedures depending on symptom distribution and surgical planning. While the majority of studies evaluated deep brain stimulation, several more recent investigations also included alternative tremor procedures such as radiofrequency thalamotomy. For example, the multi-institutional study by Goedemans (33)et al. included 35 patients treated with either DBS or radiofrequency thalamotomy for tremor. Follow-up duration varied substantially across studies, representing an additional source of heterogeneity. Prospective studies typically reported short-term outcomes at approximately three months, whereas retrospective DBS cohorts often reported longer follow-up periods extending beyond three years. Overall, the included cohorts represent clinically heterogeneous but representative populations of patients undergoing Vim-targeted surgical interventions for severe tremor.

### Targeting Strategies for Vim Localization

Across the included literature, several distinct strategies were used to localize the ventral intermediate nucleus during tremor surgery (Table 2). These approaches can broadly be categorized into four main groups: atlas-based indirect targeting, electrophysiological targeting using microelectrode recording (MER), advanced imaging-based anatomical targeting, and connectivity-based or tractography-guided targeting. In many studies, hybrid approaches combining multiple methods were used to improve targeting accuracy. Historically, the most widely used strategy has been atlas-based indirect targeting using stereotactic coordinates referenced to the AC–PC plane. Several clinical studies relied on coordinates derived from classical stereotactic atlases such as the Schaltenbrand–Wahren atlas. For example, Papavassiliou (21)et al. localized the Vim using atlas-derived coordinates relative to the midline and posterior commissure. Similarly, Chen (32)et al. performed DBS implantation using indirect stereotactic coordinates defined relative to the third ventricle and AC–PC distance. Image-guided surgical workflows have also been investigated as alternatives to electrophysiological mapping. Burchiel (14)et al. evaluated a purely imaging-guided DBS implantation technique using preoperative MRI combined with intraoperative CT verification. Despite the absence of microelectrode recording, the study demonstrated high stereotactic accuracy with a mean vector error of approximately 1.6 mm. Gravbrot (30)et al. similarly reported favorable tremor outcomes using asleep DBS procedures performed with indirect atlas targeting combined with intraoperative MRI guidance. Electrophysiological targeting using microelectrode recording has historically been considered the gold standard for functional confirmation of the Vim. This approach relies on identification of kinesthetic neurons responding to passive joint movement and evaluation of tremor suppression during intraoperative stimulation. Vassal (13)et al. described a protocol using multiple microelectrode trajectories with recordings performed in small depth increments along the distal segment of the planned trajectory. In that study, the planned central trajectory was sufficient in the majority of implantations, although additional trajectories were required in a subset of cases. More recently, several studies have explored advanced MRI techniques aimed at directly visualizing the Vim or its surrounding anatomical boundaries. Vassal et al. used WAIR MRI sequences to identify the Vim as a hypointense band within the ventrolateral thalamus. High-field imaging approaches have also been proposed. Najdenovska (23)et al. demonstrated that susceptibility-weighted imaging at 7-Tesla MRI can visualize the Vim region within the thalamus, while Su (27)et al. reported that white-matter-nulled MPRAGE imaging enables segmentation of the Vim at ultra-high field strengths. Another rapidly expanding approach involves diffusion MRI tractography targeting of the dentato-rubro-thalamic tract. Rather than targeting the Vim nucleus itself, these strategies aim to target the cerebello-thalamo-cortical pathway implicated in tremor generation. Sammartino (35)et al. demonstrated that deterministic tractography can identify the DRTT using cerebellar and cortical seed regions. Fenoy (15)et al compared tractography-guided targeting with conventional atlas-based targeting and found comparable tremor outcomes but lower stimulation requirements in the tractography-guided group. Connectivity-based targeting strategies have also been proposed to define the motor thalamus based on its structural connectivity with cortical motor regions. Middlebrooks (24)et al. used probabilistic tractography to parcellate the thalamus according to cortical connectivity patterns and demonstrated that overlap between the stimulation field and the connectivity-defined motor thalamus correlated with clinical tremor improvement. Similarly, Ferreira (31)et al. demonstrated substantial interindividual variability in connectivity-defined Vim localization, highlighting limitations of fixed stereotactic coordinates. Overall, the included studies illustrate an evolution in targeting strategies over time, progressing from traditional atlas-based coordinates toward imaging-guided and connectivity-based targeting approaches designed to enable more individualized localization of the Vim.

**Table 2.**
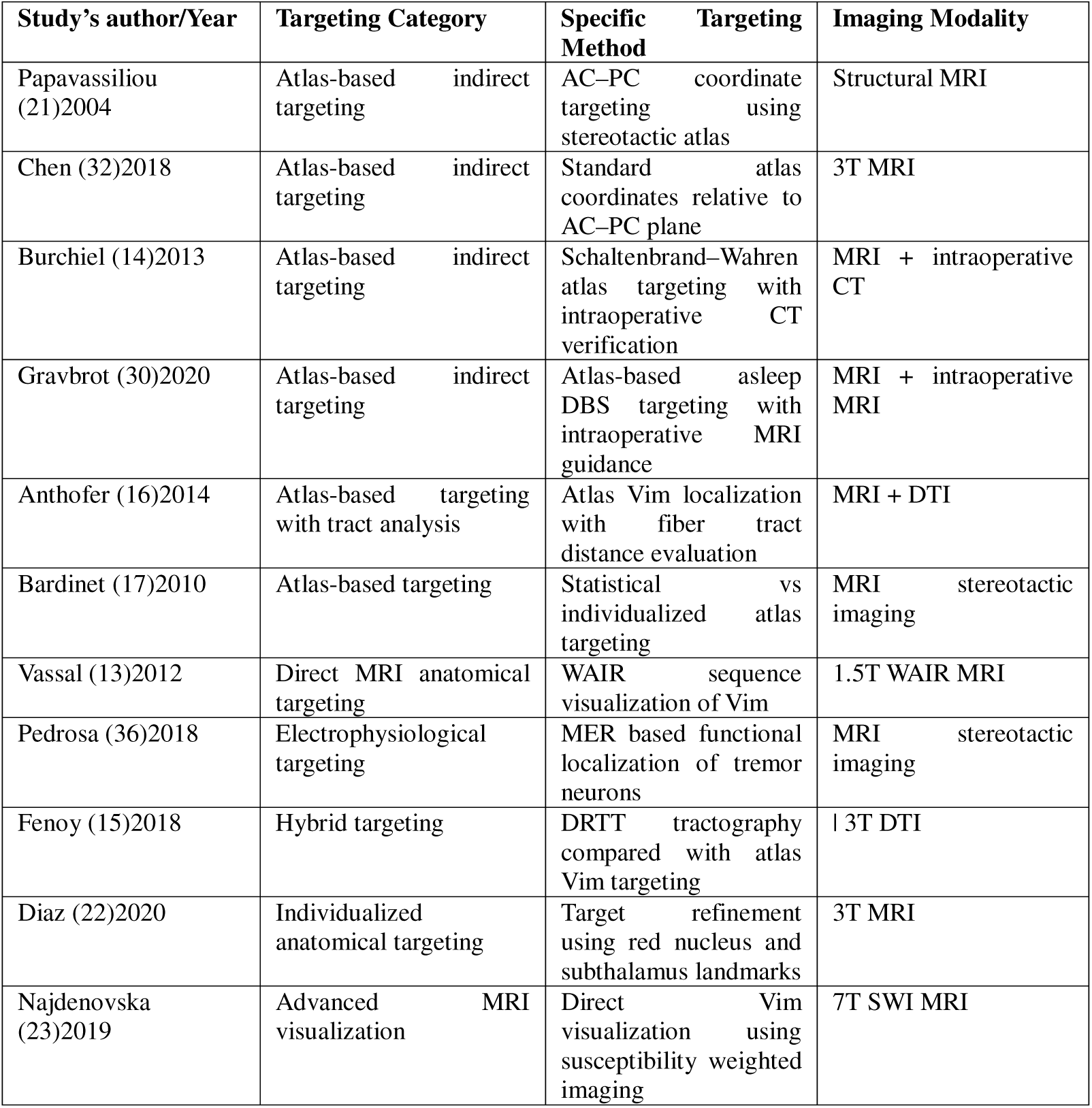
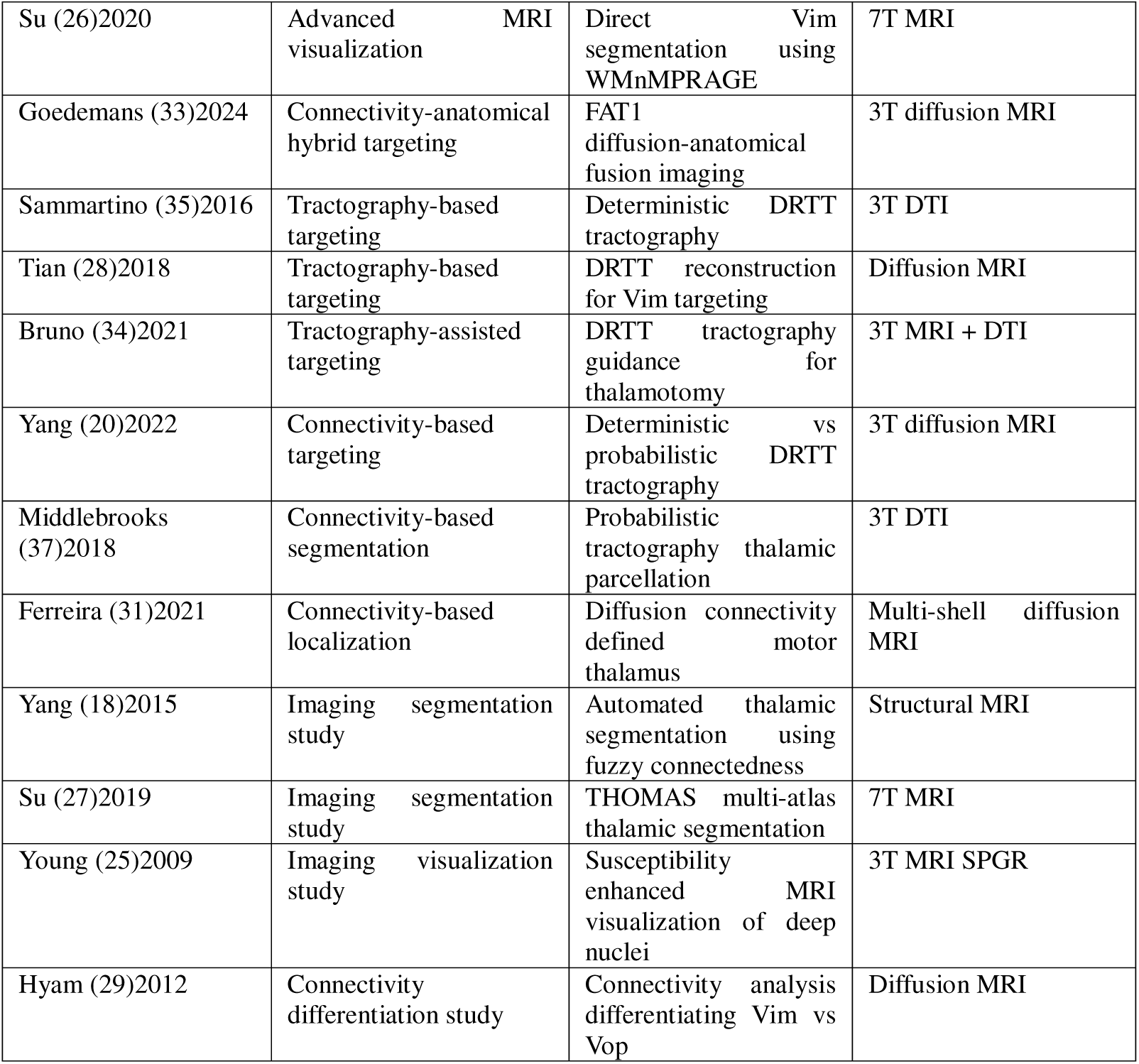
Targeting strategies used for Vim localization across included studies.

### Targeting Accuracy

Targeting accuracy was reported or could be indirectly assessed in several included studies using metrics such as vector error, radial error, Euclidean error, trajectory deviation, and the spatial distance between the planned anatomical target and the final active stimulation contact. Burchiel et al. (2013) provided one of the most detailed evaluations of stereotactic accuracy in a prospective cohort of 60 patients undergoing DBS implantation using MRI-based targeting with intraoperative CT verification. Across 119 implanted electrodes, the mean vector error between the planned target and the final electrode position was 1.59 ± 1.11 mm, and the mean deviation from the intended trajectory was 1.24 ± 0.87 mm. Targeting accuracy varied between anatomical targets, with Vim electrodes demonstrating a greater mean vector error (1.9 mm) compared with GPi electrodes (1.29 mm; p = 0.01). The authors suggested that the proximity of the Vim to the lateral ventricles may increase susceptibility to targeting deviation due to brain shift or trajectory constraints. Similarly, Chen et al. (2017) evaluated electrode placement accuracy in 56 patients undergoing Vim DBS using indirect atlas-based targeting without microelectrode recording. The mean radial error was 0.9 ± 0.3 mm in the awake group and 0.9 ± 0.4 mm in the asleep group (p = 0.75), while Euclidean error measured 1.1 ± 0.6 mm and 1.2 ± 0.5 mm, respectively (p = 0.92), demonstrating that high stereotactic precision can be achieved with image-guided techniques even without intraoperative electrophysiology. Other studies examined the relationship between anatomical targets and clinically optimal stimulation sites. Vassal et al. (2012), using direct anatomical targeting with a WAIR MRI sequence, reported a mean distance of 2.77 ± 2.1 mm between the inferior anatomical border of the Vim and the most effective stimulation contact. Despite this discrepancy, tremor improvement approached 90%, suggesting that small anatomical deviations can be compensated through intraoperative stimulation testing. Electrophysiological studies further demonstrated that functional boundaries of the Vim may extend beyond atlas-defined coordinates. Pedrosa et al. (2018) reported tremor-related neuronal activity across a depth range of approximately 5–12 mm along the stereotactic trajectory, often necessitating adjustments of final electrode placement based on physiological responses. Imaging-based targeting studies also assessed accuracy by comparing imaging-defined targets with clinically effective stimulation sites. Yang et al. (2022) reported that the mean Euclidean distance between the final active contact and the centroid of the probabilistic dentato-rubro-thalamic tract (DRTT) was 3.32 ± 1.70 mm compared with 5.01 ± 2.12 mm using deterministic tractography. Similarly, Sammartino et al. (2016) reported mean targeting errors of approximately 1.9 mm for DRTT-based targeting compared with 2.8 mm for atlas-based coordinates. Connectivity-based analyses by Ferreira et al. (2021) also demonstrated differences of approximately 1.9–2.1 mm between connectivity-defined Vim locations and traditional atlas targets. Overall, reported targeting errors across studies generally ranged between approximately 0.9 and 3 mm depending on the metric and targeting strategy used. These findings indicate that modern stereotactic techniques achieve sub-millimetric to low-millimetric precision, although the optimal functional stimulation site may not always coincide exactly with the anatomically defined Vim target.

### Imaging-Based Targeting Approaches

Substantial heterogeneity was observed across studies regarding imaging strategies used for Vim localization. Because the ventral intermediate nucleus cannot be reliably visualized using conventional MRI sequences, multiple indirect, structural, and connectivity-based approaches have been proposed. The most commonly used method remains indirect atlas-based targeting using stereotactic coordinates referenced to the AC–PC plane. Several studies in the present review used this approach based on classical stereotactic atlases such as the Schaltenbrand–Wahren atlas (Papavassiliou et al. 2004; Chen et al. 2017; Gravbrot et al. 2020; Burchiel et al. 2013). In these frameworks, the Vim target is typically located approximately 13–14 mm lateral to the midline and about 25% of the AC–PC distance anterior to the posterior commissure, with adjustments for ventricular size or thalamic morphology. However, increasing evidence suggests that substantial interindividual anatomical variability exists. Connectivity-based analyses have demonstrated that the functional motor thalamus may vary by several millimeters between individuals. For example, Ferreira et al. (2021) reported that connectivity-defined Vim locations differed from atlas-derived coordinates by up to 5 mm in some subjects, highlighting limitations of fixed stereotactic coordinates. To address these limitations, several studies have investigated advanced imaging techniques to improve direct visualization of the Vim. Vassal et al. (2012) demonstrated that a white-matter attenuated inversion recovery (WAIR) sequence at 1.5 T can identify the Vim as a hypointense band within the ventrolateral thalamus, enabling direct anatomical targeting. Ultra-high-field MRI has further improved visualization of thalamic nuclei. Najdenovska et al. (2019) reported that susceptibility-weighted imaging at 7 T can depict the Vim as a hyperintense structure within the ventrolateral thalamus, while Su et al. (2020) showed that white-matter-nulled MPRAGE imaging enables direct visualization and segmentation of the Vim. Diffusion MRI tractography has emerged as another major strategy for targeting tremor circuits by identifying the dentato-rubro-thalamic tract. Several studies have proposed targeting the thalamic segment of the DRTT rather than relying solely on atlas-based Vim coordinates (Sammartino et al. 2016; Fenoy et al 2018; Yang et al. 2022; Bruno et al. 2021). These approaches attempt to account for individual anatomical variability by identifying the center of the tremor-related pathway. Connectivity-based thalamic segmentation represents a further development of this concept. Middlebrooks et al. (2018) used probabilistic tractography to parcellate the thalamus according to cortical connectivity patterns and demonstrated that stimulation overlap with the motor thalamic segment correlated with tremor improvement after DBS. Automated segmentation techniques have also been proposed to facilitate identification of thalamic nuclei. The THOMAS multi-atlas segmentation algorithm described by Su et al. (2019) demonstrated high segmentation accuracy with Dice similarity coefficients of approximately 0.85 compared with expert manual segmentation. Overall, these studies illustrate the evolution of Vim targeting strategies from traditional atlas-based coordinates toward advanced imaging, tractography-guided targeting, and connectivity-based segmentation approaches aimed at enabling patient-specific localization of the motor thalamus.

### Risk of Bias Assessment

Methodological quality of the included studies was assessed using design-specific critical appraisal tools according to study type. Clinical observational studies, including retrospective and prospective cohort studies were evaluated using the Joanna Briggs Institute (JBI) critical appraisal instruments. (figure 2) Imaging-focused and methodological investigations were assessed using the QUADAS-2 tool. (figure 3). Eleven studies were classified as having a low risk of bias, ten as moderate risk, and four as high risk. In general, clinical cohort studies investigating surgical targeting strategies and clinical outcomes of ventral intermediate nucleus (Vim) interventions demonstrated lower risk of bias compared with imaging-methodological studies. Studies categorized as low risk of bias included Anthofer et al. (2014), Bardinet et al. (2010), Klostermann et al. (2003), Diaz et al. (2020), Middlebrooks et al. (2018), Vassal et al. (2012), Gravbrot et al. (2020), Burchiel et al. (2013), Bruno et al. (2021), and Pedrosa et al. (2018). These studies generally reported clearly defined patient populations, well-described surgical or imaging protocols, and explicit clinical outcomes such as tremor improvement or physiological confirmation of Vim localization. Moderate risk of bias was identified in several studies due to retrospective design, incomplete reporting of patient recruitment, or heterogeneity in imaging analysis pipelines. These included Yang et al. (2015), Yang et al. (2022), Papavassiliou et al. (2004), Najdenovska et al. (2019), Tian et al. (2018), Su et al. (2020), Fenoy and Schiess (2018), Chen et al. (2017), Goedemans et al. (2024), and Sammartino et al. (2016). Four studies were judged to have high risk of bias, primarily imaging methodological investigations such as Young et al. (2009), Su et al. (2019), Hyam et al. (2012), and Ferreira et al. (2021). These studies often relied on limited imaging datasets, healthy volunteer populations, or lacked clear reference standards for validating Vim localization. Across QUADAS-2 domains, the most frequent concerns involved patient selection and reference standards, whereas most JBI-assessed clinical studies demonstrated high methodological compliance despite heterogeneity in study design and imaging approaches.

**Figure 2:**
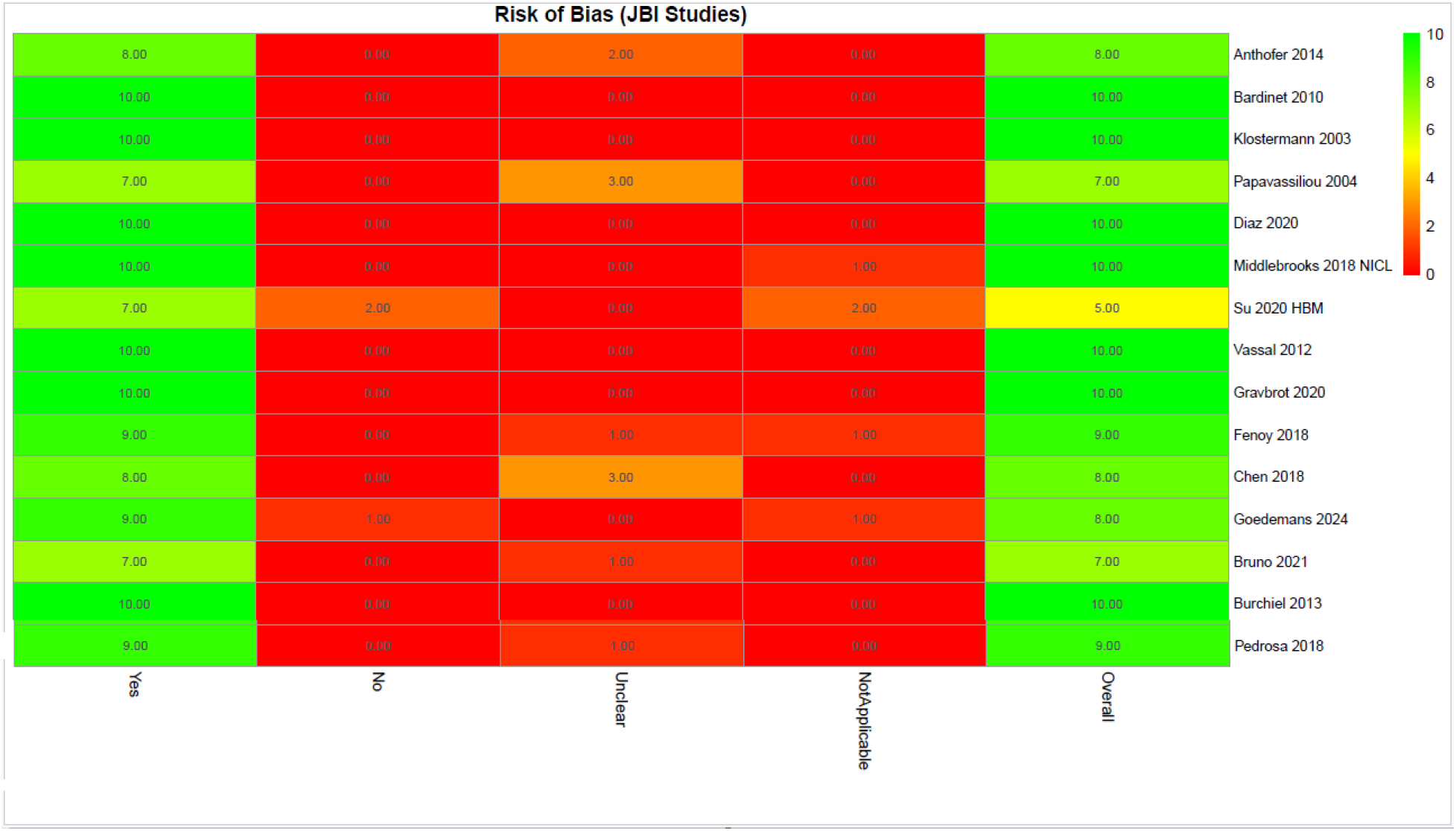
risk of bias assessment by JBI checklist.

**Figure 3:**
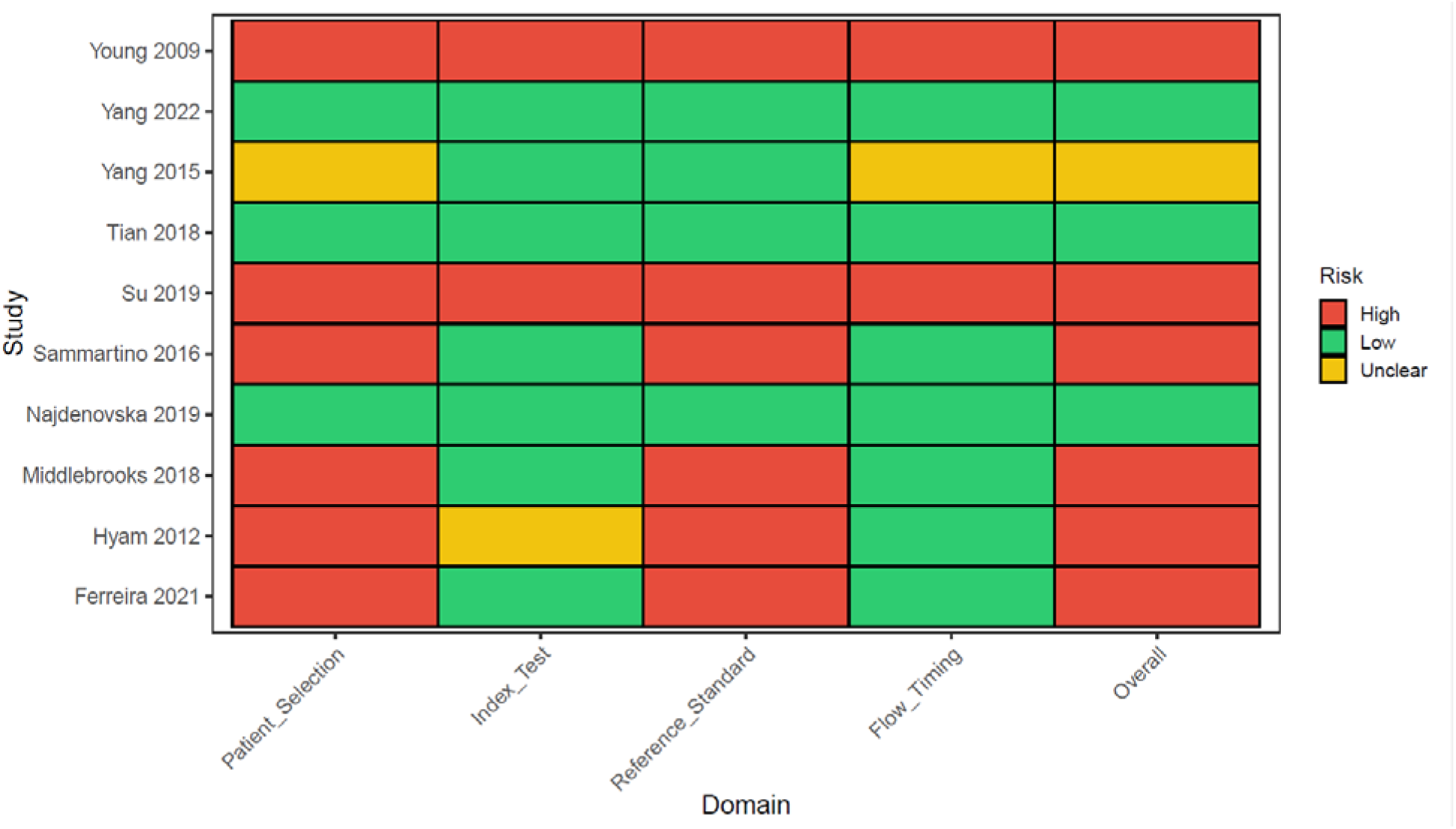
risk of bias assessment by QURADS-2 checklist.

### Eligibility for Quantitative Synthesis

Among 25 included studies for this systematic review, 7 provided sufficient quantitative outcome data from 6 studies to be included in the meta-analysis of tremor improvement. Eligibility for quantitative synthesis required studies to report extractable clinical outcomes, including pre- and postoperative tremor severity measured using validated rating scales together with sufficient statistical information to estimate measures of central tendency and variability, such as mean ± standard deviation or convertible summary statistics. Most studies were not eligible for meta-analysis because their primary objectives focused on surgical technique, targeting accuracy, electrophysiological characterization, or imaging-based methodological investigations rather than clinical tremor outcomes. Consequently, they did not report quantitative tremor rating scale data necessary for effect size calculation. For example, Burchiel et al. (2013) evaluated the stereotactic accuracy of deep brain stimulation electrode placement using MRI-based targeting with intraoperative CT verification in 60 patients. Although the study reported detailed surgical accuracy metrics, including a mean vector error of 1.59 ± 1.11 mm, it did not provide pre- or postoperative tremor severity scores and was therefore not eligible for inclusion in the meta-analysis. Similarly, Pedrosa et al. (2018) conducted an intraoperative electrophysiological mapping study of tremor-related neuronal activity in the ventrolateral thalamus but did not report standardized clinical outcome measures. In other studies, clinical improvement was reported only as percentage change without baseline or follow-up scores. For instance, Diaz et al. (2020) reported a mean tremor improvement of 79.7% ± 22.4% following individualized anatomy-based targeting, but the absence of extractable pre- and postoperative scores prevented calculation of standardized effect sizes. Additionally, some studies investigated alternative treatment modalities. Su et al. (2020), for example, evaluated imaging-based targeting for ventral intermediate nucleus ablation using magnetic resonance–guided focused ultrasound rather than DBS and did not report extractable tremor rating scores. Overall, the limited number of studies eligible for quantitative synthesis reflects the substantial methodological heterogeneity of the literature, where many investigations emphasize targeting methodology, imaging validation, or intraoperative physiology rather than standardized clinical outcome reporting.

### Pre–Post Tremor Improvement

Across the six studies included in the quantitative synthesis, comprising seven independent cohorts, tremor severity demonstrated a consistent and substantial improvement from baseline to postoperative assessment. Despite heterogeneity in study design, tremor rating scales, and anatomical targeting strategies within the ventral intermediate nucleus (Vim) region, all cohorts reported reductions in tremor severity following intervention. Standardized mean change scores were calculated from pre–post comparisons to quantify treatment effects within each cohort. Across all included datasets, standardized mean differences (SMDs) were uniformly large and negative, reflecting pronounced postoperative tremor reduction. Notably, the magnitude of improvement remained relatively consistent across studies despite the use of different clinical assessment scales, including Fahn–Tolosa–Marín Tremor Rating Scale (FTMTRS/CRST) and TETRAS-based measures. Using a random-effects model to account for between-study variability, the pooled analysis demonstrated overall treatment effect (SMD −3.91, 95% CI −4.81 to −3.01; p < 0.0001). Importantly, the confidence intervals for all individual cohorts and the pooled estimate remained entirely below zero, confirming statistically significant improvement in tremor severity across the included literature. (Figure 4)

**Figure 4.**
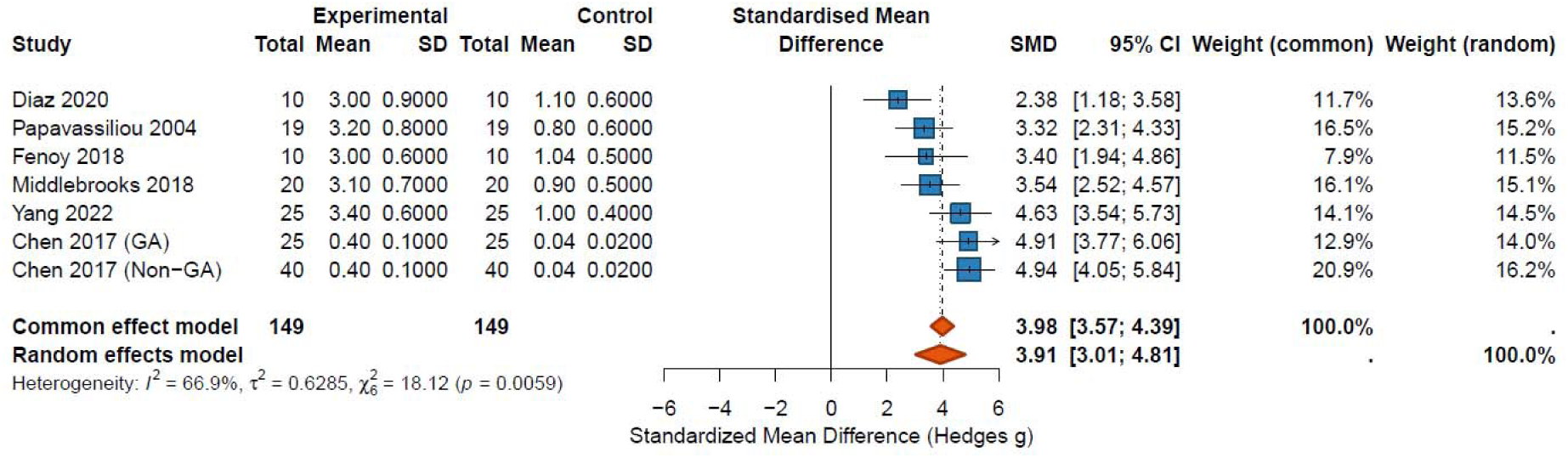
Forest plot of the meta-analysis demonstrating the pooled standardized mean difference in tremor scores between baseline and post-treatment measurements. Each square represents the effect size for an individual study, with the size of the square proportional to study weight. Horizontal lines indicate 95% confidence intervals. The diamond represents the pooled effect estimate from the random-effects model.

### Heterogeneity Analysis

Between-study heterogeneity was evaluated using Cochran’s Q statistic and the I² metric. The analysis revealed statistically significant heterogeneity (Q = 18.12, p = 0.0059), corresponding to an I² value of 66.9% and a between-study variance (τ²) of 0.63. These findings indicate that approximately two-thirds of the observed variability in effect sizes reflects true differences between studies rather than random sampling error. Several factors likely contribute to this heterogeneity, including differences in patient populations, tremor etiology, baseline tremor severity, surgical targeting approaches within the Vim complex, and variability in postoperative follow-up intervals. Nonetheless, despite these methodological and clinical differences, the direction of treatment effect remained consistent across all cohorts. To further characterize the expected variability in clinical outcomes, the random-effects model yielded a wide but consistently negative prediction interval, suggesting that future studies conducted under similar conditions would also be expected to demonstrate substantial tremor reduction following intervention.

### Sensitivity Analyses

strength of the pooled estimate was evaluated using leave-one-out sensitivity analyses. Sequential exclusion of each cohort resulted in pooled standardized mean differences ranging from −3.73 to −4.19, with all estimates remaining statistically significant. Importantly, no single study materially altered the magnitude, direction, or statistical significance of the pooled treatment effect. These findings indicate that the observed improvement in tremor severity was stable and not disproportionately driven by any individual dataset. (Figure 5)

**Figure 5.**
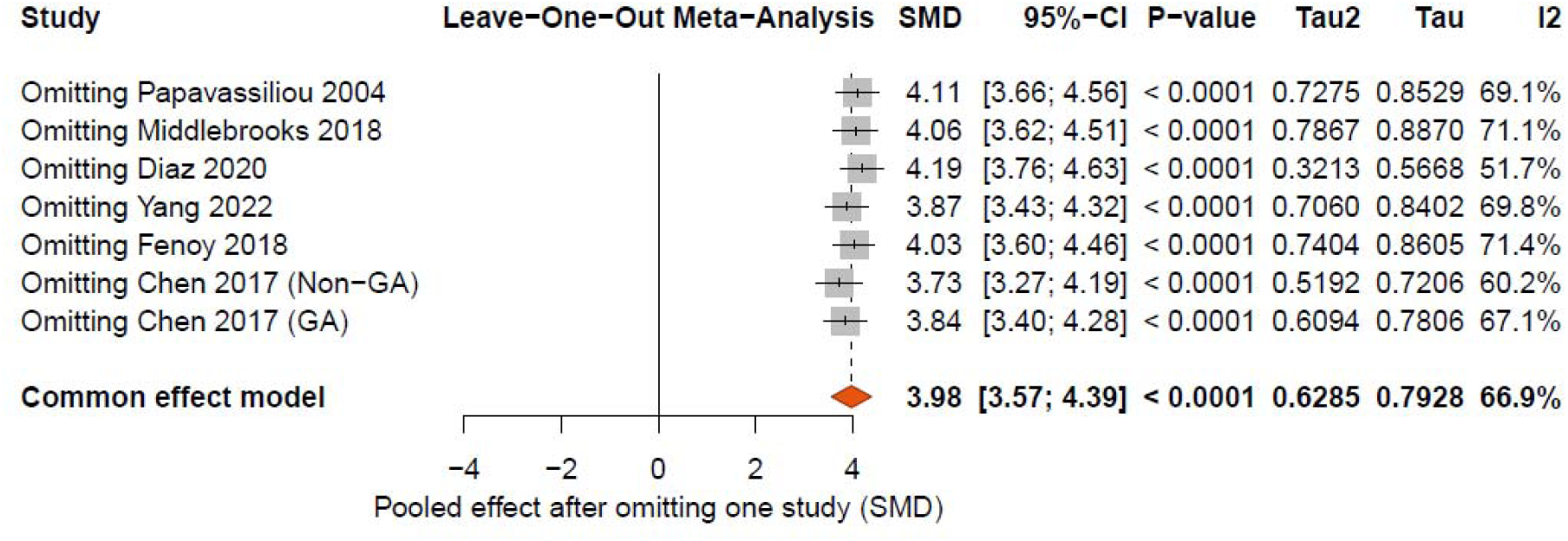
Leave-one-out sensitivity analysis showing pooled standardized mean differences after sequential removal of each study. The stability of the pooled effect size across all iterations indicates that the overall result was not driven by any single study.

### Assessment of Small-Study Effects

Potential small-study effects were explored through visual inspection of funnel plots constructed using standardized mean differences and corresponding standard errors. The distribution of effect estimates appeared broadly symmetrical around the pooled effect size, without clear evidence of asymmetry suggestive of publication bias. (Figure 8) Trim-and-fill analysis did not identify any potentially missing studies (k = 0), and the pooled effect estimate remained unchanged following adjustment. However, because the number of included studies remained below the commonly recommended threshold for reliable asymmetry testing, these findings should be interpreted cautiously. (Figure 9)

**Figure 6.**
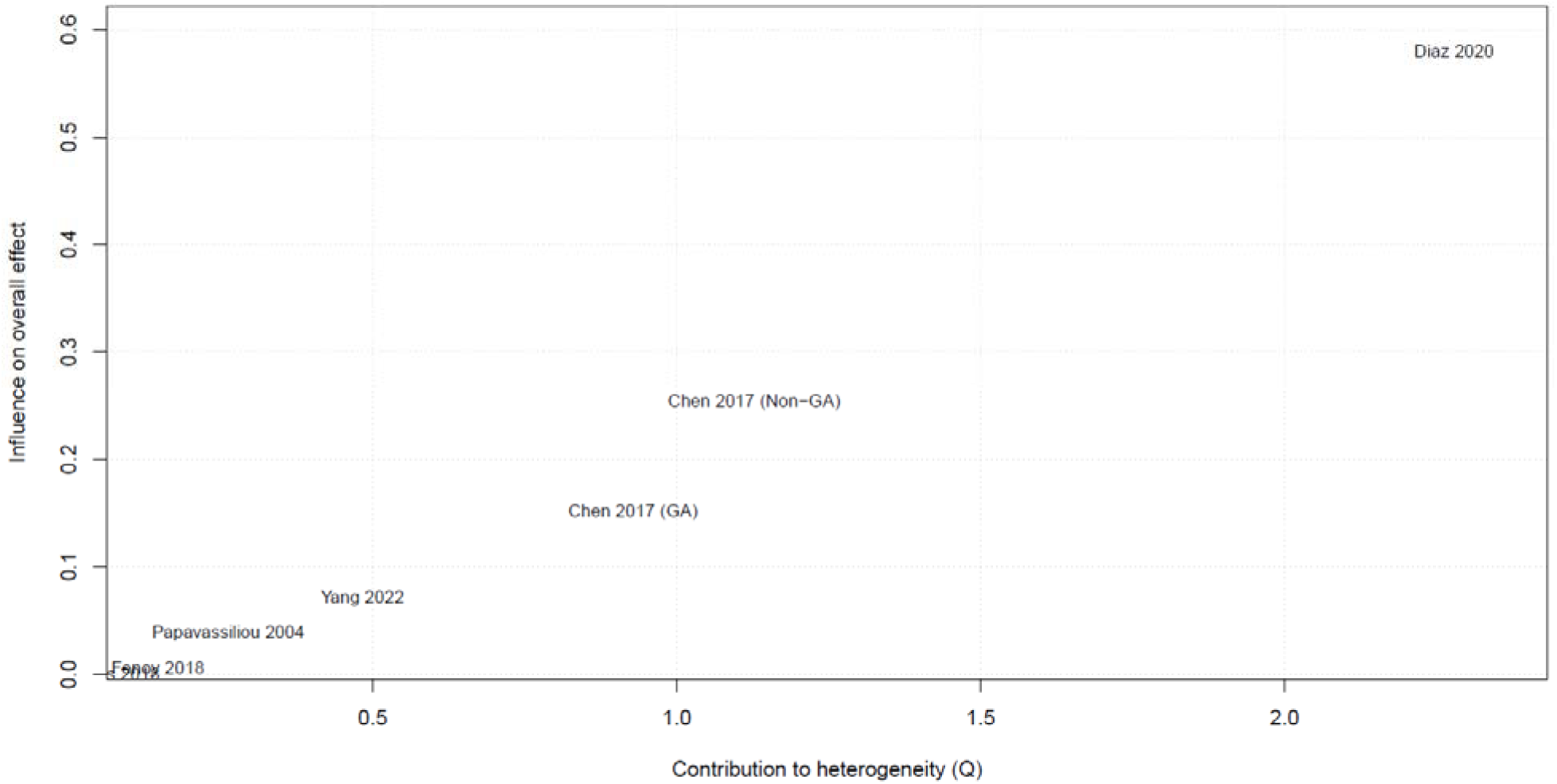
Baujat plot illustrating the contribution of each study to overall heterogeneity and influence on the pooled effect size. The study by Diaz et al. (2020) showed the largest contribution to heterogeneity but did not represent a statistical outlier.

**Figure 7.**
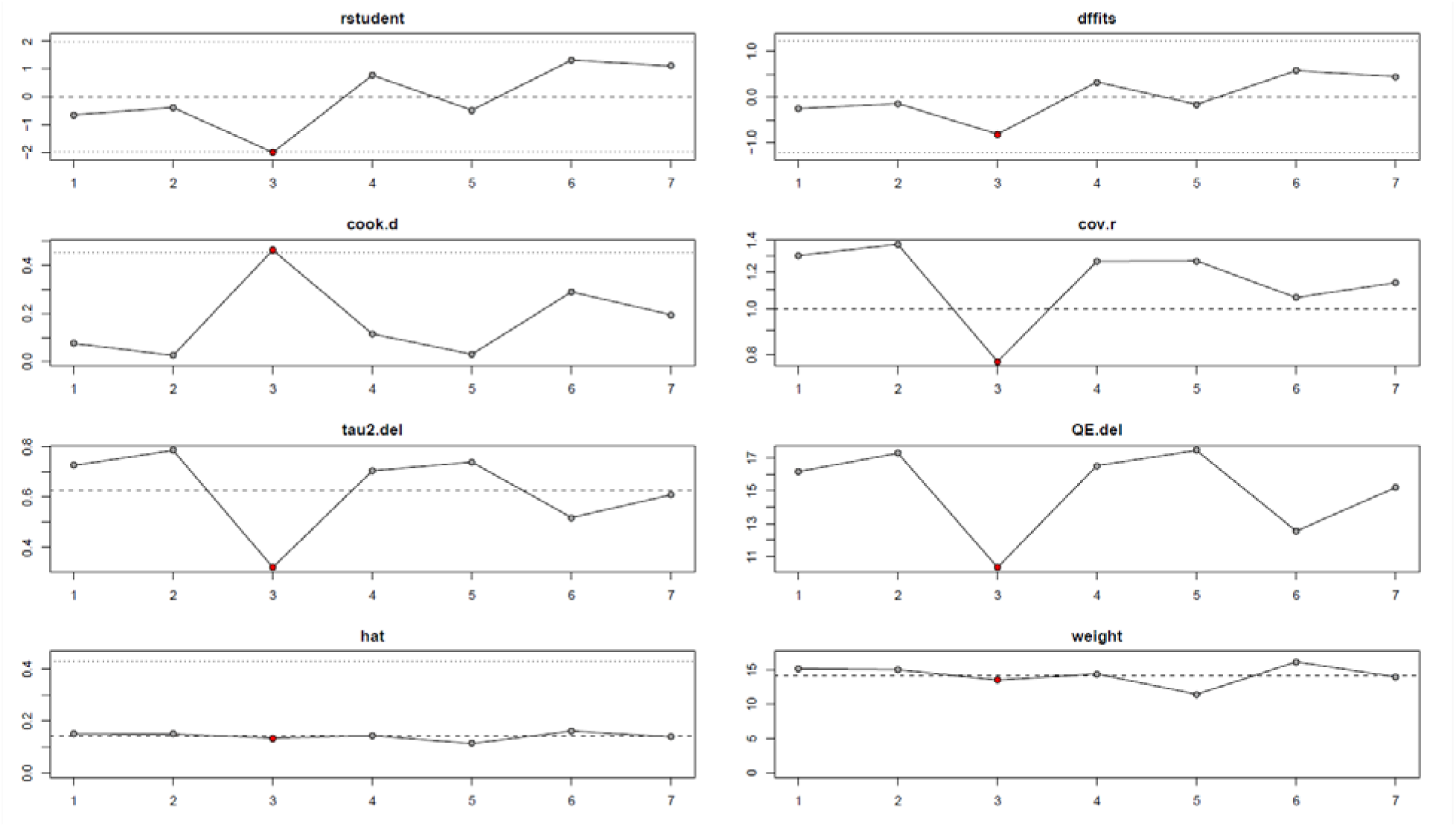
Influence diagnostics evaluating the impact of individual studies on the meta-analysis model. Metrics include studentized residuals, Cook’s distance, DFFITS, covariance ratios, hat values, and changes in heterogeneity statistics after study deletion.

**Figure 8.**
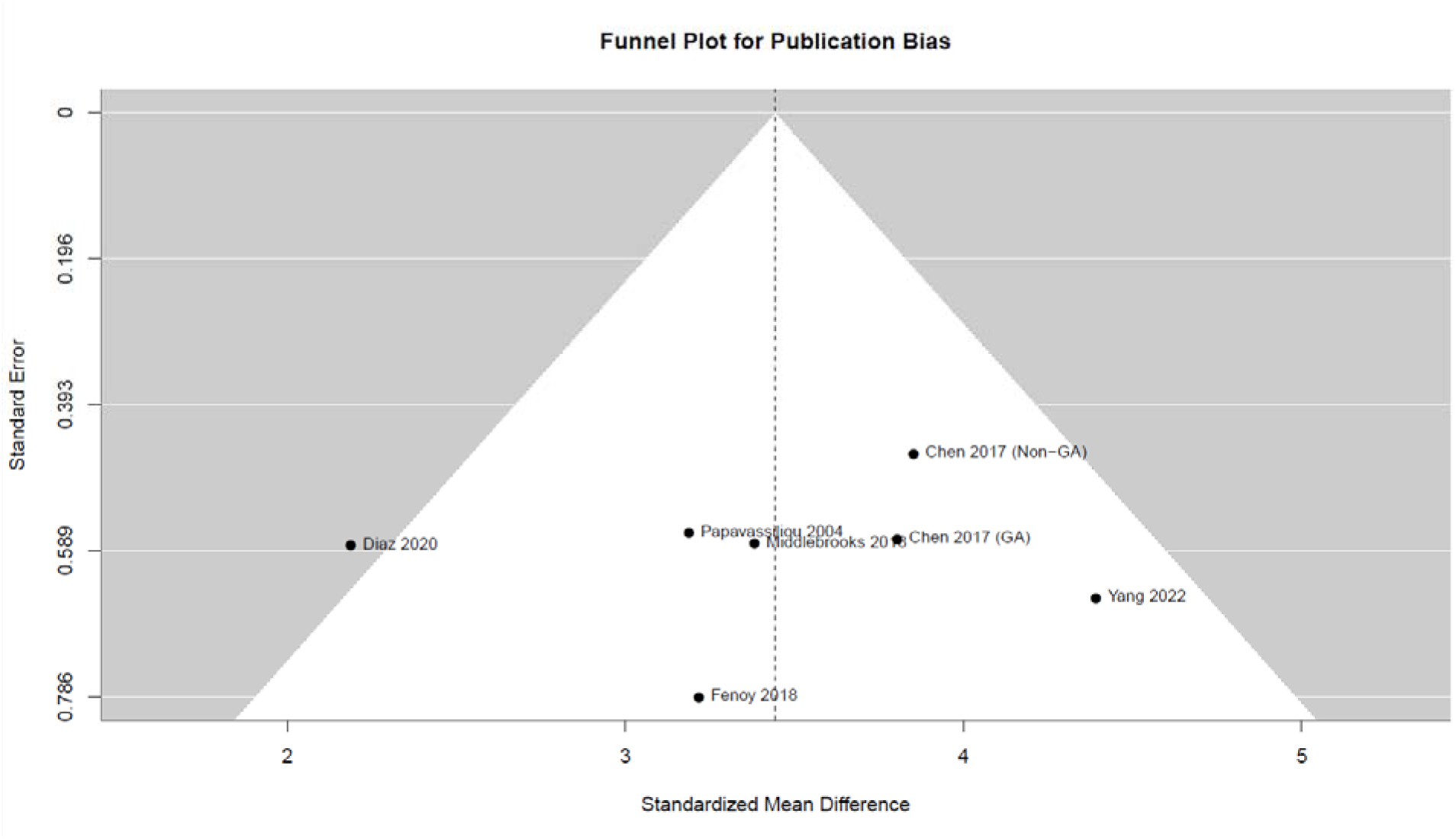
Funnel plot assessing potential publication bias in the included studies. The distribution of studies appears broadly symmetric around the pooled effect estimate.

**Figure 9.**
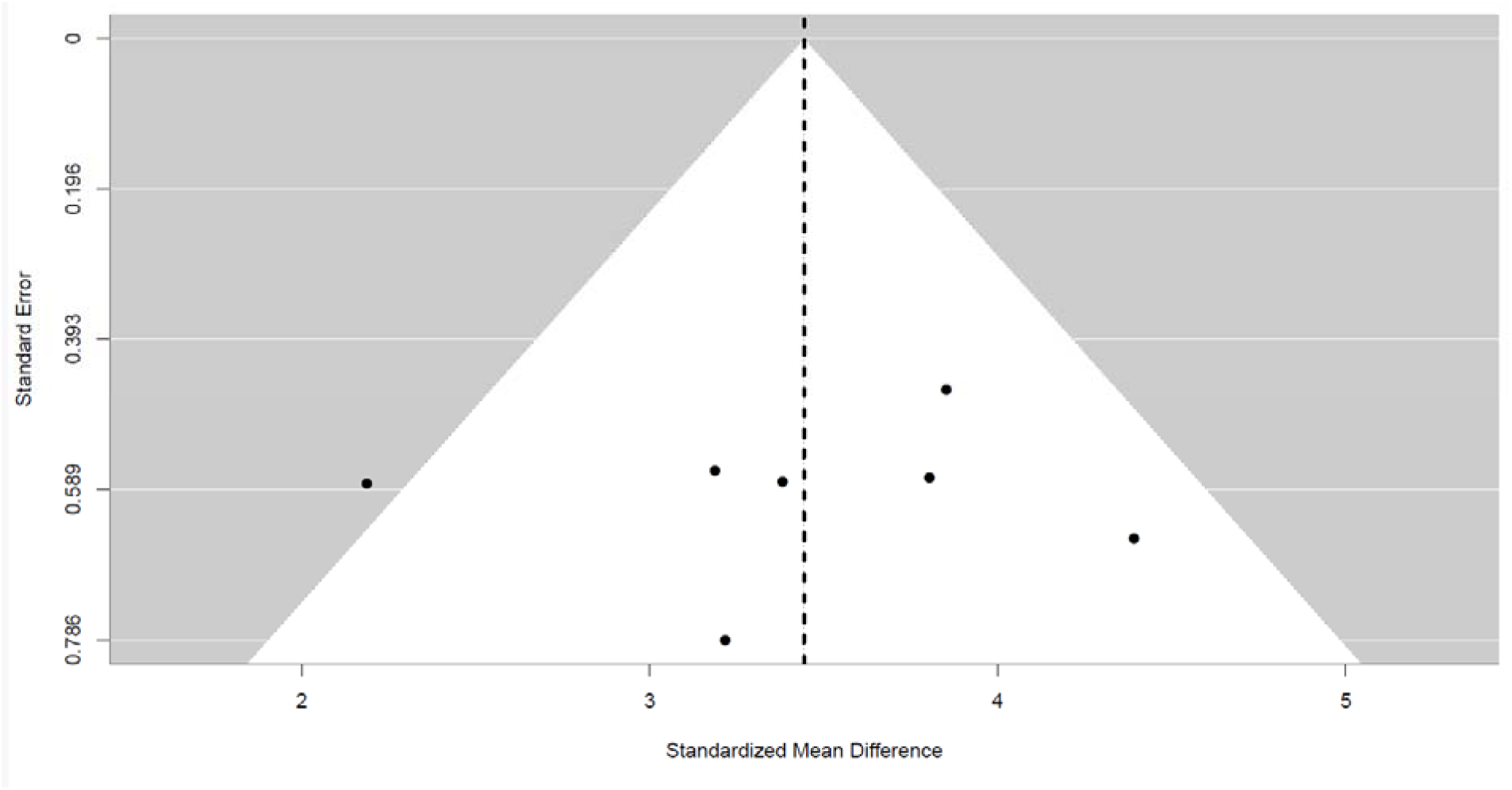
Trim and Fill plot., although with considering study number limitation, this plot did not identify any potentially missing studies (kiJ = 0).

### Influence Diagnostics

Influence diagnostics were conducted to determine whether any individual cohort exerted disproportionate influence on the pooled estimate or the observed heterogeneity. Leave-one-out analyses confirmed that removal of individual studies did not substantially alter the pooled effect size. (Figure7) Complementary evaluation using Baujat plots demonstrated that while certain cohorts contributed modestly to overall heterogeneity, no single study simultaneously exhibited high influence on the pooled estimate and high contribution to heterogeneity. (Figure6) Additional influence diagnostics, including standardized residuals, Cook’s distance, and DFFITS values, similarly indicated that none of the included cohorts exceeded conventional thresholds for undue influence. Taken together, these analyses support the robustness and internal consistency of the meta-analytic findings, reinforcing the conclusion that tremor severity improves markedly following intervention across diverse study settings and targeting methodologies.

## Discussion

### Main Findings

In this systematic review and meta-analysis, we synthesized evidence from 25 studies investigating targeting strategies for localization of the ventral intermediate nucleus (Vim) of the thalamus in tremor surgery. Across the included literature, several key findings emerged. First, the evidence demonstrates a clear methodological evolution in Vim targeting strategies, transitioning from traditional atlas-based stereotactic coordinates toward increasingly patient-specific approaches incorporating advanced MRI visualization, diffusion tractography, and connectivity-based segmentation techniques. Early studies primarily relied on indirect atlas-based coordinates relative to the anterior commissure–posterior commissure (AC–PC) plane, frequently combined with intraoperative electrophysiological confirmation using microelectrode recording (MER) and test stimulation(14, 21, 32). More recent investigations increasingly emphasize imaging-guided and connectivity-based approaches aimed at improving anatomical precision and accounting for interindividual variability in thalamic organization(20, 24, 31, 35). Second, despite differences in targeting methodology, modern stereotactic techniques appear capable of achieving consistently high spatial precision. Across studies reporting surgical accuracy metrics, targeting errors generally ranged between approximately 0.9 and 3 mm depending on the definition of accuracy and measurement method. For example, Burchiel et al. reported a mean vector error of 1.59 ± 1.11 mm for DBS electrode placement, with a slightly higher mean error of approximately 1.9 mm specifically for Vim targets(14). Similarly, Chen et al. demonstrated that purely image-guided targeting without intraoperative electrophysiological mapping achieved a mean Euclidean error of approximately 1.1 ± 0.6 mm, indicating that sub-millimetric to low-millimetric precision can be achieved using modern imaging-guided stereotactic techniques(32). Third, multiple studies highlighted an important anatomical–functional discrepancy between the anatomically defined Vim and the clinically optimal stimulation site. In a study using direct visualization of the Vim with white-matter attenuated inversion recovery (WAIR) MRI, Vassal et al. observed that the optimal stimulation contact was located on average 2.77 ± 2.1 mm away from the anatomically defined inferior Vim border, despite excellent clinical outcomes with approximately 90% tremor improvement(13). Electrophysiological mapping studies further support this concept, demonstrating that tremor-related neuronal activity can extend across several millimeters along the stereotactic trajectory and often requires intraoperative adjustments based on physiological responses(36). Fourth, emerging imaging and connectivity studies suggest that patient-specific anatomical variability may play a significant role in determining the optimal therapeutic target. Diffusion tractography investigations have shown that the centroid of the dentato-rubro-thalamic tract (DRTT), a key component of the cerebello-thalamo-cortical tremor network, may be located several millimeters away from classical atlas-based Vim coordinates. For instance, Yang et al. reported that the distance between the final active contact and the probabilistic DRTT centroid averaged 3.32 ± 1.70 mm(20), while Sammartino et al. found smaller targeting errors when using DRTT-based targeting compared with traditional atlas-based approaches (1.9 mm vs 2.8 mm)(35). Similarly, connectivity-based analyses have demonstrated that connectivity-defined Vim locations may differ from atlas-derived coordinates by approximately 1.9–2.1 mm on average and up to 5 mm in some individuals, highlighting substantial interindividual variability in the organization of the motor thalamus(31). Finally, the quantitative synthesis demonstrated robust clinical efficacy of Vim-targeted interventions for tremor control. Among the six studies (seven cohorts) providing extractable clinical outcome data, the pooled meta-analysis revealed a very large treatment effect, with a standardized mean difference of −3.91 (95% CI −4.81 to −3.01; p < 0.0001), indicating substantial postoperative reduction in tremor severity across diverse clinical settings and targeting approaches. Although moderate heterogeneity was observed across studies (I² = 66.9%), the direction of treatment effect was consistent across all cohorts, and sensitivity analyses confirmed that the overall findings were stable and not driven by any single dataset. These results suggest that Vim-based targeting strategies, despite methodological variability, consistently produce marked improvements in tremor severity in appropriately selected patients.

By integrating the available clinical outcome data across these heterogeneous studies, the present meta-analysis demonstrates a consistently large therapeutic effect associated with Vim-targeted interventions (pooled SMD −3.91, 95% CI −4.81 to −3.01), despite variability in targeting techniques, tremor rating scales, and surgical workflows. Importantly, this quantitative synthesis highlights that substantial tremor improvement is observed across diverse targeting paradigms, including atlas-based coordinates, imaging-guided approaches, tractography-assisted targeting, and electrophysiological confirmation, suggesting that effective tremor modulation may arise from engagement of the broader cerebello-thalamo-cortical network rather than precise alignment with a single fixed anatomical coordinate. Furthermore, by systematically comparing the technical accuracy and conceptual frameworks of different targeting strategies reported in the literature, this review provides a consolidated evidence base that may help guide the ongoing transition toward more individualized and connectivity-informed targeting paradigms in functional neurosurgery.

### Comparison with previous study

Several systematic reviews and meta-analyses have evaluated deep brain stimulation (DBS) across different clinical indications and stimulation targets; however, these studies have largely focused on clinical outcomes rather than the methodological aspects of ventral intermediate nucleus (VIM) localization. For example, Kinfe et al. (2026) performed an individual participant data meta-analysis examining DBS for post-stroke movement disorders and compared the efficacy of GPi, VIM, and combined GPi/VIM stimulation using standardized clinical rating scales such as the Burke–Fahn–Marsden Dystonia Rating Scale and the Fahn–Tolosa–Marín Tremor Rating Scale. Their findings suggested that pallidal stimulation was associated with significant improvement in dystonia scores, while VIM stimulation showed more limited effects; however, the study primarily addressed target selection and clinical outcomes rather than the imaging or anatomical techniques used to localize the VIM(38). Similarly, Kondapavulur et al. (2022) conducted a systematic review and network meta-analysis comparing VIM and posterior subthalamic area (PSA) stimulation for medically refractory essential tremor. Using tremor reduction measured by the Fahn–Tolosa–Marín Tremor Rating Scale as the primary outcome, their analysis demonstrated a modest short-term advantage for PSA stimulation but no significant long-term difference between targets. While this study provides important insight into comparative clinical efficacy, it was designed to evaluate tremor outcomes and did not systematically analyze the imaging protocols or targeting strategies used to identify the VIM(39). More recently, Aghajanian et al. (2025) performed a large network and multilevel meta-analysis including 52 studies comparing VIM and PSA stimulation in essential tremor. Both targets produced substantial tremor reduction, with comparable overall efficacy and a gradual decline in clinical benefit over time. Although the authors emphasized the potential importance of tractography-guided targeting and cerebello-thalamo-cortical pathways such as the dentato-rubro-thalamic tract, the analysis did not systematically examine or categorize the imaging methodologies used for VIM identification across studies(40). In addition, systematic reviews have explored DBS targets beyond the globus pallidus in specific populations. For instance, Sulistyo et al. (2025) reviewed extrapallidal DBS targets in pediatric movement disorders and identified 112 patients across 48 studies, reporting that structures such as the subthalamic nucleus and thalamic nuclei, including the VIM, have been used as alternative targets in selected cases. However, the objective of that review was to summarize clinical indications and treatment outcomes rather than to evaluate the imaging-based techniques used to localize thalamic nuclei(41). Taken together, these previous reviews provide valuable information regarding DBS target selection and clinical outcomes across different patient populations. Nevertheless, the methodological approaches used to localize the VIM, such as structural MRI visualization, atlas-based targeting, tractography-guided approaches, or segmentation strategies, have not been systematically synthesized in the existing literature. The present review therefore complements prior work by focusing specifically on the imaging and methodological aspects of VIM localization rather than on comparative clinical outcomes between stimulation targets.

### Biological Implications

The therapeutic effects of deep brain stimulation (DBS) are increasingly understood within the framework of network neuromodulation rather than modulation of a single isolated anatomical structure. In the context of tremor disorders, the ventral intermediate nucleus (VIM) of the thalamus occupies a central position within the cerebello–thalamo–cortical network, which plays a critical role in the generation and propagation of tremor-related oscillatory activity. The VIM receives afferent projections from the contralateral cerebellar dentate nucleus and relays cerebellar output to motor cortical regions, thereby functioning as a key relay node linking cerebellar and cortical motor circuits(39). Clinical and meta-analytic evidence supports the importance of this network in mediating the therapeutic effects of DBS. A systematic review and network meta-analysis comparing VIM and posterior subthalamic area (PSA) stimulation demonstrated that both targets produce substantial tremor reduction in patients with essential tremor, although PSA stimulation may offer modest short-term advantages in some cohorts (39). Importantly, the PSA region contains cerebellothalamic fibers of the dentato-rubro-thalamic tract (DRTT), suggesting that modulation of this pathway may represent a shared mechanistic substrate underlying tremor suppression. Similarly, a more recent network and multilevel meta-analysis including 52 studies confirmed that both VIM and PSA stimulation produce clinically meaningful tremor reduction with comparable overall efficacy, further highlighting the central role of cerebellothalamic pathways in tremor pathophysiology(40). Additional evidence from other movement disorder populations reinforces the concept that tremor and related motor symptoms arise from dysfunction within distributed neural circuits. An individual participant data meta-analysis evaluating DBS for post-stroke movement disorders reported heterogeneous responses depending on the stimulation target, with globus pallidus interna (GPi) stimulation demonstrating greater improvements in dystonia, whereas VIM stimulation showed more variable effects in tremor-dominant phenotypes(38). These findings underscore the functional specialization of different nodes within the motor network and suggest that optimal neuromodulation strategies depend on the underlying circuit pathology. Beyond classical movement disorders, DBS studies across different neurological conditions further support the concept of network-based neuromodulation. For instance, a systematic review of extrapallidal DBS targets in pediatric movement disorders identified the VIM as one of the most frequently used thalamic targets outside the globus pallidus, particularly in patients with tremor and dystonia, highlighting its broader role within motor control circuits(41). Similarly, meta-analytic evidence indicates that DBS can significantly reduce tremor severity in patients with multiple sclerosis–related tremor, where the VIM is commonly targeted as part of the cerebellothalamic pathway involved in tremor generation(42). The network-based mechanism of DBS is also illustrated in disorders affecting non-motor neural circuits. For example, stimulation of the fornix in patients with Alzheimer’s disease has been proposed as a strategy to modulate the Papez memory circuit and hippocampal connectivity, reflecting the broader principle that DBS can influence distributed neural networks involved in cognitive function(4). Similarly, meta-analytic comparisons of subthalamic nucleus (STN) and globus pallidus internus (GPi) stimulation in Parkinson’s disease demonstrate that modulation of distinct nodes within the basal ganglia–thalamo–cortical network can lead to comparable improvements in tremor control(3)

Taken together, these findings highlight that the therapeutic effects of DBS are best understood within the framework of large-scale neural networks. Within this context, the VIM functions as a critical relay node in the cerebello–thalamo–cortical pathway, and its stimulation likely disrupts pathological oscillatory activity propagated through cerebellothalamic circuits such as the dentato-rubro-thalamic tract. Improved characterization of these structural and functional pathways through advanced neuroimaging and tractography techniques may therefore enhance target localization and optimize DBS outcomes. Consequently, understanding the neurobiological architecture of the VIM and its associated networks remains essential for refining imaging-based targeting strategies and improving therapeutic precision in DBS.

## Clinical Implications

The findings of this review have several practical implications for surgical decision-making during deep brain stimulation (DBS) for tremor disorders. In patients undergoing DBS under awake or lightly sedated conditions, intraoperative assessment of tremor can provide valuable physiological confirmation of accurate ventral intermediate nucleus (VIM) targeting. Because the VIM functions as a relay within the cerebello–thalamo–cortical network implicated in tremor generation, intraoperative tremor suppression observed during test stimulation may serve as an important functional indicator of appropriate lead placement. Evidence from prior clinical and meta-analytic studies has demonstrated that stimulation of the VIM can produce clinically meaningful tremor reduction in patients with movement disorders, supporting its role as a key neuromodulation target. This consideration becomes particularly relevant in clinical environments where access to advanced neuroimaging technologies may be limited. Modern targeting strategies increasingly rely on high-resolution structural imaging such as 3-Tesla or 7-Tesla MRI, diffusion tractography, and emerging artificial intelligence–based targeting methods. However, these technologies are not universally available and may impose significant financial and infrastructural burdens in many centers. In such settings, physiological confirmation through intraoperative tremor evaluation can serve as a practical adjunct to anatomical targeting, allowing surgeons to refine lead positioning even in the absence of advanced imaging resources. From a procedural perspective, performing DBS implantation under awake or sedated conditions enables real-time clinical testing and physiological confirmation of target engagement. In many institutions, when this approach is used, implantation of the implantable pulse generator (IPG) in the subpectoral or pectoral region is subsequently performed during a second surgical stage. This staged strategy remains common practice when DBS is not performed entirely under general anesthesia and allows the surgical team to prioritize optimal electrode positioning during the initial implantation procedure. Importantly, the findings of the present study also suggest that the absence of high-cost imaging technologies should not necessarily preclude the feasibility of VIM-DBS surgery. In resource-limited environments, the potential need for a staged surgical approach may provide a practical pathway to circumvent technological limitations. Accepting a two-stage surgical workflow, in which physiological intraoperative testing is used to guide electrode placement and the pulse generator is implanted during a later procedure, may help overcome the lack of advanced imaging infrastructure and alleviate concerns that DBS surgery cannot be safely performed without such technologies. Conversely, in patients who require general anesthesia, DBS implantation is frequently performed as a single-stage procedure, where intraoperative clinical testing is not feasible. In these cases, alternative strategies may be required to approximate physiological targeting. Imaging-based simulation approaches or preoperative virtual targeting techniques derived from structural and diffusion imaging may therefore serve as useful adjuncts to guide lead placement when intraoperative feedback cannot be obtained. Taken together, these considerations highlight the importance of integrating anatomical imaging, physiological feedback, and surgical workflow considerations when planning DBS procedures. Tailoring the operative strategy according to patient characteristics, available technology, and institutional resources may ultimately improve targeting precision.

## Future Directions

Future research should focus on improving the precision and reproducibility of ventral intermediate nucleus (VIM) targeting in deep brain stimulation (DBS) surgery. First, further development and validation of preoperative imaging-based targeting strategies are required. Advanced neuroimaging techniques, including high-resolution structural MRI and diffusion tractography, may enable more accurate visualization of cerebello–thalamo–cortical pathways implicated in tremor generation. Incorporating multimodal imaging into preoperative planning may therefore enhance anatomical localization of the VIM and reduce variability in electrode placement across surgical centers.

Second, emerging neuromodulation technologies may open new avenues for selective targeting of tremor-related neural circuits. In particular, nanoparticle-based approaches may represent a novel strategy for delivering targeted neuromodulatory agents or enabling localized modulation of deep brain structures. Although such techniques remain largely experimental, both preclinical and translational investigations could explore whether nanoparticle-mediated targeting of the VIM or its associated pathways may provide complementary or alternative therapeutic strategies for tremor modulation.

Third, the integration of physiological biomarkers during intraoperative targeting represents another promising direction. Tremor disorders are characterized by abnormal oscillatory activity within the cerebello–thalamo–cortical network. The development of higher-resolution electroencephalography (EEG) systems or other advanced neurophysiological monitoring techniques may enable more precise detection of tremor-related oscillatory signals during DBS procedures. Incorporating these physiological markers into the targeting process could potentially improve functional confirmation of optimal stimulation sites and reduce reliance on expensive imaging technologies.

Fourth, additional efforts are needed to develop standardized anatomical and functional atlases that accurately define the boundaries of the VIM and its associated fiber pathways. Current literature demonstrates variability in anatomical definitions and targeting strategies, which complicates comparisons across studies and may contribute to inconsistent surgical outcomes. High-resolution probabilistic atlases integrating structural, diffusion, and functional imaging data may help establish more consistent targeting frameworks for DBS surgery.

Finally, future imaging and electrophysiological studies should carefully consider the clinical heterogeneity of patients undergoing VIM-DBS for treatment-resistant tremor. Many of these patients may present with comorbid neurological conditions, structural brain lesions, or disease-related neuroanatomical alterations that can influence both brain anatomy and physiological network activity. Prior studies have highlighted substantial heterogeneity and potential selection bias in DBS populations, which limits the strength of current evidence and complicates interpretation of outcomes (Kinfe et al., 2026). Consequently, future investigations should prioritize larger sample sizes and study designs that account for patient-specific anatomical and physiological characteristics. Developing personalized targeting strategies that integrate clinical, imaging, and neurophysiological data may ultimately improve the precision of VIM localization and optimize therapeutic outcomes in DBS surgery.

## Limitations

Several limitations should be considered when interpreting the findings of this systematic review and meta-analysis. First, although 25 studies met the eligibility criteria for inclusion in the systematic review, only four studies provided sufficient quantitative data on preoperative and postoperative tremor outcomes to be included in the meta-analysis. The limited number of studies eligible for quantitative synthesis reduces the statistical power of the pooled analysis and restricts the ability to perform more robust subgroup or sensitivity analyses.

Second, the included literature demonstrated substantial methodological heterogeneity across multiple domains. The studies varied widely in their design, including retrospective cohorts, prospective observational studies, methodological imaging investigations, and case series. As described in the included dataset, several studies primarily focused on imaging validation or anatomical localization of the Vim rather than reporting standardized clinical outcomes. This variability in study objectives and methodologies complicates direct comparison across studies and may influence the generalizability of the pooled findings.

Third, patient populations were heterogeneous across the included studies. Although most cohorts involved patients with medically refractory tremor, the underlying etiologies differed considerably, including essential tremor, Parkinson’s disease tremor, mixed tremor syndromes, and in some cases datasets including healthy participants for imaging validation. Such variability in disease populations may introduce clinical heterogeneity that could influence both targeting strategies and treatment outcomes.

Fourth, many of the included studies were based on relatively small sample sizes. As shown in the study characteristics table, several investigations included fewer than 20 participants, and some imaging methodological studies involved even smaller datasets. Small cohort sizes may increase the risk of statistical instability and limit the external validity of the reported findings.

Fifth, heterogeneity was also observed in the methods used to localize the ventral intermediate nucleus. Targeting approaches included indirect atlas-based stereotactic coordinates, direct MRI visualization techniques, tractography-based targeting of the dentato-rubro-thalamic tract, connectivity-based thalamic segmentation, and physiological confirmation using microelectrode recording. The coexistence of these diverse targeting strategies reflects the evolving landscape of Vim localization but also limits the ability to draw definitive conclusions regarding the relative superiority of specific techniques.

Sixth, outcome reporting across studies was inconsistent. Tremor severity was assessed using different clinical rating scales, including the Fahn–Tolosa–Marin Tremor Rating Scale (FTMTRS), the Essential Tremor Rating Assessment Scale (TETRAS), and other global clinical outcome measures. The lack of a uniform outcome metric introduces additional variability and may affect the comparability of treatment effects across studies included in the quantitative synthesis.

Seventh, several potentially relevant studies could not be incorporated into the meta-analysis due to insufficient quantitative reporting of outcomes. The absence of standardized numerical data in these reports limited the possibility of including them in the pooled analysis and may have resulted in the exclusion of valuable clinical information.

Eighth, the small number of studies included in the quantitative analysis limited the reliability of formal publication bias assessment. Although funnel plot analysis was attempted, the limited dataset restricts the interpretability of such analyses, and potential publication bias cannot be definitively excluded.

Finally, risk-of-bias assessment indicated that several included studies had moderate to high risk of bias, largely due to retrospective study designs, limited sample sizes, and variability in reporting of surgical techniques and outcome measures. These factors should be considered when interpreting the overall conclusions of the present study.

## CONCLUSION

The present systematic review demonstrates that the literature on ventral intermediate nucleus targeting for tremor is characterized by substantial methodological diversity, encompassing atlas-based stereotactic approaches, electrophysiological mapping with microelectrode recordings, advanced structural MRI techniques, and connectivity-based tractography strategies. Despite these differences in anatomical localization methods, the available clinical evidence consistently shows marked postoperative improvement in tremor severity following intervention. The quantitative synthesis confirmed a large and statistically significant reduction in tremor scores across the included cohorts, with pooled effect estimates demonstrating treatment efficacy. Although moderate heterogeneity was observed, likely reflecting variations in patient populations, tremor etiologies, targeting strategies, and follow-up intervals, the direction of treatment effect remained uniform across all studies. Sensitivity and influence analyses further supported the stability of the pooled findings. These results suggest that clinically tremor suppression can be achieved across a range of Vim targeting methodologies. While emerging imaging and connectivity-based techniques may improve anatomical precision and contribute to a more individualized surgical approach, current evidence does not yet establish clear superiority of any single targeting strategy in terms of clinical tremor outcomes. Future research should focus on prospective comparative studies with standardized tremor rating scales, uniform reporting of targeting coordinates and imaging parameters, and longer-term follow-up. Such efforts will be essential to determine whether modern imaging-guided targeting approaches translate into measurable improvements in surgical accuracy, clinical outcomes, and long-term durability of tremor control.

## Supporting information

data extraction

full text exclusion

full text inclusion

full result

checklist prisma

protocol prospero

risk of bias

search strategy

## Data Availability

All data produced in the present study are available upon reasonable request to the authors

## Declaration of AI Use

Artificial intelligence (AI) tools (ChatGPT) were used to improve the grammar, clarity, and language of this manuscript. The authors reviewed and approved all content, and take full responsibility for the integrity and accuracy of the work.

## Acknowledgments

The authors have no acknowledgments to declare.

## Appendix and Supplementary Material

- Appendix 1. PRISMA checklist
- Appendix 2. Prospero Protocol
- Appendix 3. Search Strategy
- Appendix 4. Title-Abstract screening excluded studies
- Appendix 5,6. Full text screening Excluded and Included studies
- Appendix 7. Data extraction Sheets
- Appendix 8. Risk of bias
- Appendix 9. Full Result Version

## Ethics Approval and Consent to Participate

This study is a systematic review and meta-analysis based exclusively on previously published studies. No primary data were collected and no direct involvement of human participants occurred. Therefore, ethical approval from an Institutional Review Board (IRB) or Ethics Committee was not required.

## Consent to Participate declaration in the manuscript

Not applicable.

## Human Ethics and Consent to Participate declarations

Not applicable.

## Clinical trial number

not applicable.

## Author Contributions

Farzan Fahim conceived and supervised the study, designed the methodological framework, and developed the standardized screening and data extraction forms. He oversaw the entire review process, prepared the study selection documentation, verified extracted datasets, resolved reviewer disagreements, and provided final approval of the analytical dataset and manuscript. MohammadAmin Farajzadeh contributed to study coordination and methodological calibration. He performed the pilot calibration step for the screening process, randomly selected studies for inter-reviewer agreement assessment, and conducted training sessions for reviewers involved in the risk-of-bias evaluation procedures. Sadra Abedinzadeh and Deniz Pourkhalil independently conducted the title and abstract screening and participated in full-text eligibility assessment according to the predefined inclusion and exclusion criteria. Deniz Pourkhalil and Mojtaba Esmaeeli independently performed data extraction from all eligible studies using the standardized extraction forms developed for the project. Extracted datasets were subsequently reviewed and validated under the supervision of the senior author. Salar Pirbabaee and Fatemeh Salarifar independently performed the risk-of-bias assessment for all included studies using the Joanna Briggs Institute (JBI) Critical Appraisal Checklist for Cohort Studies and the QUADAS-2 tool for imaging and diagnostic methodology studies, following methodological training. authors contributed to interpretation of the findings, critical revision of the manuscript for important intellectual content, and approval of the final version of the manuscript prior to submission.

