## Supplementary material for "Tremor Improvement Despite Heterogeneous Ventral Intermediate Nucleus Targeting in Deep Brain Stimulation: A Systematic Review and Meta-Analysis": full result

RESULTS

**Study Selection**

The study selection process is summarized in the PRISMA flow diagram(fig.1). A total of 2,398 records were identified through database searching, with no additional records retrieved from registers. After removal of 1,016 duplicate records, 1,382 studies remained for title and abstract screening. During screening, 1,235 records were excluded for the following reasons: conference abstracts, protocols, or registry-only reports (n = 70), studies not relevant to the predefined PICOS criteria (n = 570), and studies with an inappropriate design or population (n = 595). A total of 147 full-text reports were sought for retrieval, of which 7 could not be obtained. Consequently, 140 full-text articles were assessed for eligibility. Among these, several studies were excluded for the following reasons: case series with fewer than 10 participants (n = 11), studies targeting nuclei other than the ventral intermediate nucleus (Vim) (n = 50), studies employing irrelevant methodological approaches (n = 28), studies not specifically addressing Vim targeting (n = 20), and studies with ineligible designs (n = 6). Ultimately, 25 studies met the eligibility criteria and were included in the systematic review. These consisted of 7 methodological or imaging studies, 13 retrospective cohort studies, 3 prospective cohort studies, 1 case series, and 1 observational cross-sectional study. Among these, four studies provided sufficient quantitative data on preoperative and postoperative tremor scores and were therefore included in the quantitative meta-analysis.

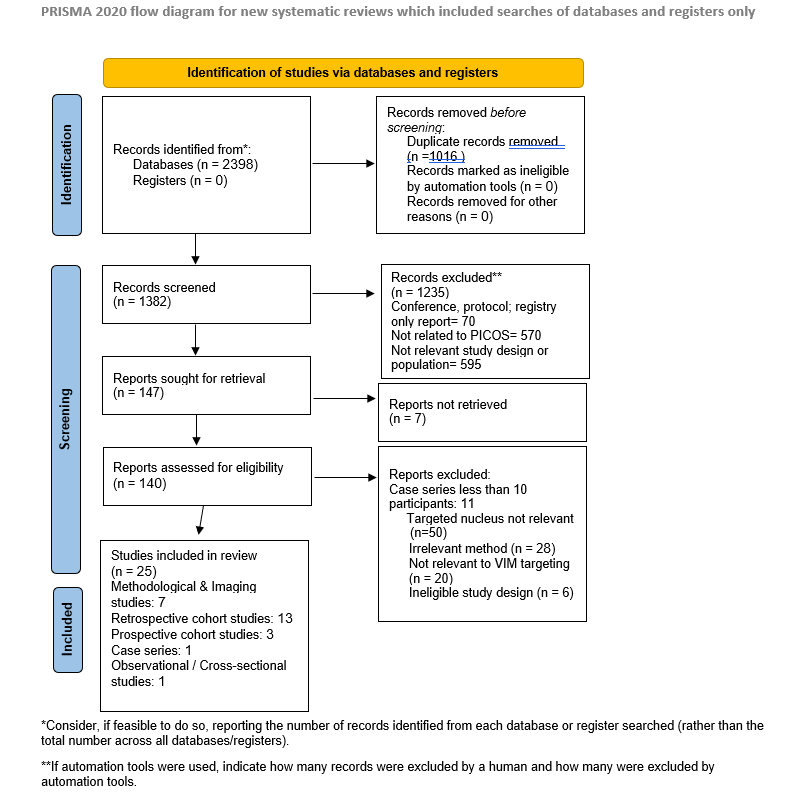

**Study Characteristics**

The included studies demonstrated substantial methodological heterogeneity in study design, patient populations, imaging strategies, and approaches used for localization and validation of the ventral intermediate nucleus (Vim) of the thalamus. Across the included literature, study designs comprised retrospective case series, retrospective observational cohorts with post‑hoc imaging analyses, prospective cohort studies, and comparative cohort analyses evaluating different targeting strategies. For example, Vassal et al. (2012) conducted a retrospective consecutive case series of patients undergoing Vim deep brain stimulation (DBS) for medically refractory tremor, whereas Burchiel et al. (2013) performed a prospective cohort study assessing the surgical accuracy of image‑guided DBS electrode placement using intraoperative computed tomography. Other studies adopted comparative methodological designs, such as Fenoy and Schiess (2018), who compared tractography‑assisted targeting of the dentato‑rubro‑thalamic tract (DRTt) with conventional atlas‑based stereotactic targeting of the Vim. Patient populations were also heterogeneous across studies. Most cohorts consisted of patients with medically refractory tremor syndromes, primarily essential tremor (ET) and Parkinson’s disease tremor, although the relative proportion of each diagnosis varied between studies. For instance, the cohort reported by Vassal et al. (2012) included both Parkinson’s disease and essential tremor patients undergoing Vim DBS implantation, while Fenoy et al. (2018) focused specifically on patients with essential tremor. In contrast, the multi‑institutional imaging study by Goedemans et al. (2024) included mixed tremor etiologies and combined treatment modalities, including both DBS and radiofrequency thalamotomy. Substantial variability was also observed in targeting methodologies. Several studies relied on conventional indirect stereotactic targeting based on anterior commissure–posterior commissure (AC–PC) coordinates and stereotactic atlases such as the Schaltenbrand–Bailey or Morel atlas. However, multiple investigations evaluated advanced imaging‑based targeting strategies aimed at improving visualization of the intrathalamic anatomy. For example, Vassal et al. (2012) used a white‑matter‑attenuated inversion recovery (WAIR) MRI sequence at 1.5‑T to directly visualize the Vim as a hypointense band within the ventrolateral thalamus. In contrast, Burchiel et al. (2013) implemented a purely image‑guided surgical workflow combining preoperative 3‑T MRI with intraoperative CT verification of electrode placement, without the use of microelectrode recording (MER). More recent studies have explored connectivity‑based or diffusion‑guided approaches. Fenoy et al. (2018) used deterministic diffusion tractography to directly target the dentato‑rubro‑thalamic tract, representing a network‑based targeting strategy rather than direct anatomical Vim localization. Similarly, Goedemans et al. (2024) evaluated an advanced imaging technique termed FAT1 imaging, which combines fractional anisotropy diffusion maps with T1‑weighted MRI to enhance visualization of intrathalamic nuclei and improve targeting accuracy. Intraoperative validation techniques also varied across studies. Some surgical workflows incorporated microelectrode recording to physiologically confirm the Vim through identification of kinesthetic neurons responsive to passive joint movement and tremor‑related activity, as described in the protocol used by Vassal et al. (2012). Conversely, other studies intentionally omitted electrophysiological mapping in favor of imaging‑based verification of electrode placement, as in the CT‑guided workflow reported by Burchiel et al. (2013). In addition, several studies used postoperative imaging analyses to quantify targeting accuracy or overlap between stimulation volumes and imaging‑defined targets. Outcome reporting across studies was likewise heterogeneous. Clinical tremor outcomes were assessed using a variety of rating scales, including the Fahn–Tolosa–Marin Tremor Rating Scale (FTMTRS), the Essential Tremor Rating Assessment Scale (TETRAS), or global clinical improvement measures. Follow‑up duration varied substantially between studies, ranging from short‑term follow‑up of a few months to long‑term clinical assessments exceeding three years. Overall, the included literature reflects the evolving landscape of Vim targeting strategies in tremor surgery, progressing from traditional atlas‑based stereotactic coordinates toward advanced imaging‑guided and connectivity‑based targeting methods. However, the substantial heterogeneity in study design, patient populations, imaging protocols, validation techniques, and outcome measures should be considered when interpreting the findings across studies. summary of included studies provided in Table1.

| **First author/Year/ country** | **Study design** | **ample Size** | **Disease Population** | **Targeting Strategy** | **Imaging Method** | **Follow‑up** | **Main Clinical Outcome** |
| --- | --- | --- | --- | --- | --- | --- | --- |
| Anthofer 2014 / Germany | Retrospective observational | 10 | ET, PD, dystonia, OCD | Atlas‑based indirect targeting | MRI + DTI tractography | Not reported | Fiber tract distance analysis relative to atlas Vim |
| Bardinet 2010 \| France | Prospective observational | 29 | Tremor | Statistical vs individualized atlas targeting | MRI stereotactic imaging | 1–6 months | 80% tremor improvement |
| Yang 2015 \| China | Imaging methodological | 25 | Imaging dataset | Automated thalamic segmentation | Structural MRI | Not Applicable | Dice similarity coefficient 0.88 |
| Klostermann 2003 \| Germany | Imaging methodological | 23 | PD, ET, MS tremor | SEP‑based physiological localization | MRI + CT fusion | Immediate postop | 60% tremor improvement |
| Yang 2022 \| USA | Retrospective cohort | 19 | ET | Probabilistic vs deterministic DRTT targeting | 3T DTI tractography | 3.1 months | 66% tremor improvement |
| Papavassiliou 2004 \| USA | Retrospective cohort | 37 | ET | Postoperative MRI correlation of lead location | MRI structural imaging | 26 months | 53% tremor reduction |
| Diaz 2020 \| USA | Retrospective cohort | 33 | ET | Individualized anatomy‑based targeting | 3T MRI anatomical landmarks | 31 months | 79.7% tremor improvement |
| Najdenovska 2019 \| Switzerland | Imaging methodological | 11 | Healthy + ET \| | Direct visualization on 7T SWI | 7T MRI | 3–9 months imaging follow‑up | Validation of Vim visualization |
| Middlebrooks 2018 \| USA | Retrospective cohort | 40 | ET | Connectivity‑based thalamic segmentation | 3T DTI probabilistic tractography | 6 months | 41% tremor improvement |
| Young 2009 \| USA | Imaging methodological | 4 | Healthy volunteers | Susceptibility‑enhanced MRI targeting | 3T MRI SPGR | Not Applicable | Visualization of midbrain nuclei |
| Su 2020 \| USA | Retrospective cohort | 14 | ET | Direct visualization on 7T WMnMPRAGE | 7T MRI | 1 month | 65.7% tremor improvement |
| Su 2019 \| USA | Imaging methodological | 35 | Mixed population | Multi‑atlas segmentation (THOMAS) | 7T MRI | Not Applicable | Dice coefficient 0.85 |
| Vassal 2012 \| France | Retrospective cohort | 20 | PD, ET | Direct MRI visualization (WAIR) | 1.5T WAIR MRI | 46 months | 90% tremor improvement |
| Tian 2018 \| USA | Retrospective cohort | 15 | ET | DRTT tractography targeting | 3T diffusion MRI | 3–12 months | 68–76% tremor improvement |
| Hyam 2012 \| UK | Imaging methodological | 17 | Healthy volunteers | Connectivity differentiation Vim vs Vop | 5T DTI | Not Applicable | Connectivity differences demonstrated |
| Gravbrot 2020 \| USA | Case series | 12 | ET | Atlas‑based indirect targeting (asleep DBS) | MRI + intraoperative MRI | 6 months | 71% tremor improvement |
| Ferreira 2021 \| UK | Imaging methodological | 100 | Healthy controls | Connectivity‑defined Vim localization | 3T multi‑shell diffusion MRI | Not Applicable | Demonstrated atlas variability |
| Fenoy 2018 \| USA | Retrospective cohort | 40 | ET | DRTT tractography vs atlas targeting | 3T DTI | 12 months | 69% vs 62% improvement |
| Chen 2017 \| USA | Prospective cohort | 56 | ET | Indirect atlas targeting | 3T MRI | 3 months | 48% ADL improvement |
| Goedemans 2024 \| UK | Prospective cohort | 35 | Tremor (ET, others) | FAT1 diffusion‑anatomical fusion imaging | 3T MRI | 13 months | 41.7% favorable outcome |
| Bruno 2021 \| Italy | Retrospective cohort | 21 | ET, PD tremor | Tractography vs atlas targeting | 3T MRI + DTI | 6 months | 63% tremor reduction |
| Sammartino 2016 \| Canada | Retrospective cohort | 14 | ET | DRTT tractography targeting | 3T DTI | 6 months | 62% tremor reduction |
| Pedrosa 2018 \| Germany | Retrospective cohort | 10 | ET | MER‑based physiological targeting | MRI stereotactic | Not reported | Functional tremor suppression |
| Anthofer 2014 \| Germany | Retrospective cohort | 10 | ET, PD | Atlas targeting with tract distance analysis | MRI + DTI | Not reported | Spatial variability assessment |
| Yang 2015 \| China | Imaging methodological | 25 | Imaging dataset | Fuzzy connectedness segmentation | MRI | Not Applicable | Automated thalamic segmentation validation |

Table 1. summary of included studies.

Abbreviations: ADL: Activities of Daily Living /CGI‑I: Clinical Global Impression–Improvement scale /cZI: Caudal zona incerta /DRTT: Dentato‑rubro‑thalamic tract /ET: Essential tremor /FA: Fractional anisotropy /FAT1: Fusion of Fractional Anisotropy maps with T1‑weighted anatomical MRI /FTMTRS: Fahn–Tolosa–Marin Tremor Rating Scale /HARDI: High angular resolution diffusion imaging/ HCP: Human Connectome Project /MER: Microelectrode recording /MNI: Montreal Neurological Institute stereotactic space /MPRAGE: Magnetization‑prepared rapid gradient echo/ MS: Multiple sclerosis /OCD: Obsessive‑compulsive disorder /PD: Parkinson’s disease /PLR: Prelemniscal radiations /RF‑T: Radiofrequency thalamotomy /ROI: Region of interest /SPGR: Spoiled gradient recalled /TETRAS: The Essential Tremor Rating Assessment Scale /THOMAS: Thalamus Optimized Multi‑Atlas Segmentation /UPDRS: Unified Parkinson’s Disease Rating Scale /VC: Ventrocaudal nucleus of the thalamus /Vop: Ventral oral posterior nucleus of the thalamus /VTA: Volume of tissue activated/ WAIR: White matter attenuated inversion recovery

**Patient Characteristics**

Across the included studies, a total of 211 patients were analyzed. The study populations primarily consisted of individuals undergoing surgical treatment for medically refractory tremor syndromes. Eligibility criteria were generally comparable across studies and typically required disabling tremor despite adequate pharmacological therapy. Essential tremor represented the most common underlying diagnosis across cohorts, although several studies also included patients with Parkinson disease tremor. For example, in the study by Vassal et al. (2012), 20 patients with medically refractory tremor underwent Vim deep brain stimulation (DBS), including 13 patients with Parkinson disease tremor and 7 patients with essential tremor. Similarly, the prospective cohort reported by Burchiel et al. (2013) included 60 patients undergoing DBS for movement disorders, comprising Parkinson disease (n = 33), essential tremor (n = 26), and dystonia (n = 1); among these, 25 patients underwent DBS targeting of the ventral intermediate nucleus (Vim). Several studies focused exclusively on essential tremor populations. Chen et al. (2017) evaluated 56 patients with essential tremor undergoing Vim DBS implantation and compared two surgical approaches: awake DBS surgery (n = 16) and asleep DBS surgery under general anesthesia (n = 40). Similarly, Fenoy and Schiess (2018) conducted a comparative cohort study of 40 patients with essential tremor who underwent DBS implantation using either conventional atlas‑based Vim targeting (n = 20) or tractography‑assisted targeting of the dentato‑rubro‑thalamic tract (n = 20). Demographic characteristics were broadly similar across studies, with most cohorts consisting of older adults. The reported mean age ranged from approximately 61.7 ± 15.8 years to 67.7 ± 8.8 years. For instance, the mean age in the cohort reported by Vassal et al. was 62.5 ± 15.5 years, while Burchiel et al. reported a mean age of 64 ± 9.5 years. In the study by Chen et al., the mean age was 61.7 ± 15.8 years in the awake DBS group and 67.7 ± 8.8 years in the asleep DBS group. Sex distribution, when reported, showed a predominance of male patients; for example, 70% of participants were male in the Vassal et al. cohort. The majority of surgical interventions involved deep brain stimulation targeting the ventral intermediate nucleus of the thalamus. Both unilateral and bilateral implantation strategies were used across studies. In the Vassal et al. cohort, bilateral implantation was performed in 12 patients and unilateral implantation in 8 patients, resulting in a total of 32 implanted thalami. In the Chen et al. study, a combination of unilateral and bilateral procedures resulted in 90 implanted electrodes. More recent imaging‑focused studies also included alternative tremor surgeries. For example, the multi‑institutional cohort reported by Goedemans et al. (2024) included 35 patients (43 treated surgical sides), of whom 16 underwent DBS implantation and 19 underwent radiofrequency thalamotomy. In this cohort, essential tremor accounted for 66% of cases and Parkinson disease tremor for 34%, with a mean age of 66 ± 8 years and a predominance of male patients (66%). Follow‑up durations varied substantially across studies, representing an additional source of clinical heterogeneity. Prospective cohorts such as Chen et al. reported short‑term outcomes at approximately three months, whereas retrospective DBS cohorts reported longer follow‑up periods. For example, Vassal et al. reported a mean follow‑up duration of 46.3 ± 28.7 months (range 8–86 months). Overall, the included studies represent heterogeneous but clinically representative cohorts of patients undergoing Vim‑targeted neurosurgical interventions for severe tremor refractory to medical therapy.

**Targeting Strategies for Vim Localization**

Across the 25 studies included in this systematic review, several distinct surgical targeting strategies were used for localization of the ventral intermediate nucleus (Vim) during tremor surgery which summarized in Table 2. These approaches can be broadly categorized into four main strategies: (1) atlas‑based indirect targeting, (2) intraoperative electrophysiological targeting using microelectrode recording (MER) or test stimulation, (3) advanced imaging‑based anatomical targeting, and (4) connectivity‑based or tractography‑guided targeting. Many studies employed hybrid approaches that combined two or more of these strategies to compensate for the limitations of any single method.

The most widely used approach historically is indirect atlas‑based targeting based on stereotactic coordinates referenced to the anterior commissure–posterior commissure (AC–PC) plane. Several clinical DBS studies in the present review relied primarily on this strategy, using stereotactic atlases such as the Schaltenbrand–Wahren atlas or Guiot coordinates to define the Vim target relative to midline structures. For example, Papavassiliou et al. (2004) localized the Vim approximately 12.3 mm lateral to the midline and 6.3 mm anterior to the posterior commissure using atlas‑derived coordinates. Similarly, Chen et al. (2017) performed Vim DBS using indirect targeting defined as approximately 10.5 mm lateral to the third ventricular wall (maximum ≈14 mm from the midline) and approximately 25% of the AC–PC distance posterior to the mid‑commissural point. In this prospective cohort of 56 essential tremor patients, both awake DBS and asleep DBS procedures used the same atlas‑based coordinates and achieved comparable clinical outcomes, with approximately 48% improvement in tremor‑related activities of daily living (Chen et al., 2017).

Atlas‑based targeting was also used in several studies evaluating image‑guided DBS implantation without electrophysiological mapping. In the prospective study by Burchiel et al. (2013), electrode placement was performed using MRI‑based anatomical targeting combined with intraoperative CT verification without microelectrode recording. The targeting coordinates for Vim were derived from the Schaltenbrand and Wahren atlas relative to the AC–PC plane. Despite the absence of MER, the study demonstrated high stereotactic accuracy with a mean vector error of 1.59 ± 1.11 mm across implanted electrodes, supporting the feasibility of purely imaging‑guided DBS implantation in selected patients (Burchiel et al., 2013). Similarly, Gravbrot et al. (2020) reported that asleep DBS performed using indirect atlas‑based targeting combined with interventional MRI guidance achieved tremor improvements of approximately 71% with mean radial placement error of approximately 0.5 mm, suggesting that modern imaging guidance may compensate for the absence of intraoperative electrophysiology.

A second major targeting strategy involves intraoperative electrophysiological mapping, particularly microelectrode recording (MER) and intraoperative test stimulation. This approach is designed to identify the functional boundaries of the Vim by recording neuronal activity and evaluating clinical tremor suppression during stimulation. Electrophysiological mapping has historically been considered the gold standard for functional target confirmation because the Vim cannot be reliably visualized on conventional MRI. Several studies included in this review employed MER to refine electrode placement. For example, Vassal et al. (2012) used two exploratory microelectrode trajectories per hemisphere, with recordings performed along the distal 10 mm of the planned trajectory in 0.5‑mm steps. Kinesthetic neurons responding to passive joint movement were used as physiological markers of the Vim, and intraoperative macrostimulation was applied to evaluate tremor arrest and side‑effect thresholds. In this cohort of 20 patients, the central planned trajectory was sufficient in approximately 84.5% of cases, while additional trajectory adjustments were required in approximately 15.5% of implantations (Vassal et al., 2012).

Similarly, Pedrosa et al. (2018) demonstrated that MER can identify tremor‑related neuronal activity within the ventral thalamus across multiple recording depths, providing functional confirmation of the target prior to final electrode implantation. In that study, tremor‑modulated neuronal activity was recorded along trajectories extending 5–12 mm around the planned stereotactic target, highlighting the functional complexity of the motor thalamus and the potential limitations of relying solely on anatomical coordinates. In comparative surgical workflows, MER is often combined with intraoperative test stimulation to identify stimulation thresholds associated with tremor suppression and stimulation‑induced adverse effects such as paresthesia or dysarthria (Fenoy and Schiess, 2018).

More recently, several studies have explored advanced imaging‑based targeting strategies designed to directly visualize the Vim or its anatomical boundaries. Because conventional MRI sequences often fail to delineate thalamic subnuclei, specialized imaging sequences have been developed to improve thalamic contrast. One of the earliest approaches was described by Vassal et al. (2012), who used a white matter attenuated inversion recovery (WAIR) sequence at 1.5 T to directly identify the Vim as a hypointense band within the ventrolateral thalamus. This structure was bordered posteriorly by the ventrocaudal nucleus and inferiorly by the prelemniscal radiations, allowing direct anatomical targeting rather than relying solely on atlas‑based coordinates. Using this imaging approach combined with intraoperative confirmation, the authors reported approximately 90% improvement in tremor severity at long‑term follow‑up (Vassal et al., 2012).

High‑field MRI techniques have also been investigated for direct visualization of the Vim. Najdenovska et al. (2019) demonstrated that susceptibility‑weighted imaging at 7‑Tesla MRI can identify the Vim as a hyperintense structure within the ventrolateral thalamus. Similarly, Su et al. (2020) reported that white‑matter‑nulled MPRAGE imaging allows clear visualization and segmentation of the Vim at ultra‑high field strengths, enabling patient‑specific targeting for MR‑guided focused ultrasound procedures. More recently, Goedemans et al. (2024) introduced a hybrid imaging technique combining diffusion‑derived fractional anisotropy with T1‑weighted anatomical imaging (FAT1 imaging), which allows visualization of the motor thalamus and Vim in a single structural image dataset. In that study involving 35 tremor patients treated with DBS or radiofrequency thalamotomy, the FAT1‑defined target demonstrated strong correlation with clinical outcomes, suggesting that diffusion‑anatomical fusion imaging may represent a promising future approach for patient‑specific targeting.

Another rapidly expanding targeting strategy involves diffusion MRI tractography to identify the dentato‑rubro‑thalamic tract (DRTT), a key cerebello‑thalamo‑cortical pathway implicated in tremor generation. Rather than targeting the Vim nucleus directly, these approaches aim to target the thalamic segment of the DRTT or its intersection with the motor thalamus. Sammartino et al. (2016) demonstrated that deterministic tractography can identify the DRTT using seed regions in the cerebellar dentate nucleus and termination regions in the motor cortex. Using this strategy, the authors reported that tractography‑defined targets were closer to the clinically effective stimulation site compared with traditional atlas‑based coordinates. Similarly, Fenoy and Schiess (2018) compared direct DRTT targeting with conventional atlas‑based Vim targeting in a cohort of 40 patients with essential tremor. Both approaches produced significant tremor reduction at 1‑year follow‑up; however, the tractography‑guided group required significantly lower stimulation frequencies, suggesting improved stimulation efficiency.

Additional studies have further supported the potential advantages of connectivity‑based targeting strategies. Yang et al. (2022) compared deterministic and probabilistic tractography methods for identifying the DRTT and demonstrated that probabilistic tractography produced targets that were spatially closer to the final active DBS contact. Similarly, Bruno et al. (2021) evaluated tractography‑guided targeting during MR‑guided focused ultrasound thalamotomy and found that tractography‑derived targets closely approximated clinically effective ablation sites, often requiring only small spatial adjustments during treatment planning.

Finally, several studies have explored connectivity‑based thalamic segmentation techniques designed to define the Vim based on its structural connectivity to motor cortical regions. Middlebrooks et al. (2018) used probabilistic tractography to parcellate the thalamus according to its connectivity with cortical motor regions, demonstrating that stimulation overlap with the motor thalamic segment correlated with tremor improvement following DBS. Similarly, Ferreira et al. (2021) demonstrated substantial interindividual variability in the location of connectivity‑defined Vim regions, with atlas‑based coordinates differing by several millimeters in some individuals. These findings further emphasize the limitations of fixed stereotactic coordinates and support the development of patient‑specific targeting strategies.

Taken together, the studies included in this review illustrate a clear evolution in Vim targeting strategies over time. Early DBS procedures relied predominantly on atlas‑based coordinates combined with electrophysiological confirmation. More recent approaches increasingly incorporate advanced MRI visualization, diffusion tractography, and connectivity‑based segmentation to achieve patient‑specific targeting. While no single targeting strategy has emerged as universally superior, the available evidence suggests that combining high‑resolution imaging with functional or connectivity‑based information may provide the most accurate and reproducible localization of the Vim for tremor surgery.

| **Study’s author/Year** | **Targeting Category** | **Specific Targeting Method** | **Imaging Modality** |
| --- | --- | --- | --- |
| Papavassiliou 2004 | Atlas‑based indirect targeting | AC–PC coordinate targeting using stereotactic atlas | Structural MRI |
| Chen 2017 | Atlas‑based indirect targeting | Standard atlas coordinates relative to AC–PC plane | 3T MRI |
| Burchiel 2013 | Atlas‑based indirect targeting | Schaltenbrand–Wahren atlas targeting with intraoperative CT verification | MRI + intraoperative CT |
| Gravbrot 2020 | Atlas‑based indirect targeting | Atlas‑based asleep DBS targeting with intraoperative MRI guidance | MRI + intraoperative MRI |
| Anthofer 2014 | Atlas‑based targeting with tract analysis | Atlas Vim localization with fiber tract distance evaluation | MRI + DTI |
| Bardinet 2010 | Atlas‑based targeting | Statistical vs individualized atlas targeting | MRI stereotactic imaging |
| Vassal 2012 | Direct MRI anatomical targeting | WAIR sequence visualization of Vim | 1.5T WAIR MRI |
| Pedrosa 2018 | Electrophysiological targeting | MER based functional localization of tremor neurons | MRI stereotactic imaging |
| Fenoy 2018 | Hybrid targeting | DRTT tractography compared with atlas Vim targeting | \| 3T DTI |
| Diaz 2020 | Individualized anatomical targeting | Target refinement using red nucleus and subthalamus landmarks | 3T MRI |
| Najdenovska 2019 | Advanced MRI visualization | Direct Vim visualization using susceptibility weighted imaging | 7T SWI MRI |
| Su 2020 | Advanced MRI visualization | Direct Vim segmentation using WMnMPRAGE | 7T MRI |
| Goedemans 2024 \| \| \| \| | Connectivity‑anatomical hybrid targeting | FAT1 diffusion‑anatomical fusion imaging | 3T diffusion MRI |
| Sammartino 2016 | Tractography‑based targeting | Deterministic DRTT tractography | 3T DTI |
| Tian 2018 | Tractography‑based targeting | DRTT reconstruction for Vim targeting | Diffusion MRI |
| Bruno 2021 | Tractography‑assisted targeting | DRTT tractography guidance for thalamotomy | 3T MRI + DTI |
| Yang 2022 | Connectivity‑based targeting | Deterministic vs probabilistic DRTT tractography | 3T diffusion MRI |
| Middlebrooks 2018 | Connectivity‑based segmentation | Probabilistic tractography thalamic parcellation | 3T DTI |
| Ferreira 2021 | Connectivity‑based localization | Diffusion connectivity defined motor thalamus | Multi‑shell diffusion MRI |
| Yang 2015 | Imaging segmentation study | Automated thalamic segmentation using fuzzy connectedness | Structural MRI |
| Su 2019 | Imaging segmentation study | THOMAS multi‑atlas thalamic segmentation | 7T MRI |
| Young 2009 | Imaging visualization study | Susceptibility enhanced MRI visualization of deep nuclei | 3T MRI SPGR |
| Hyam 2012 | Connectivity differentiation study | Connectivity analysis differentiating Vim vs Vop | Diffusion MRI |

Table 2. Targeting strategies used for Vim localization across included studies

**Targeting Accuracy**

Targeting accuracy was explicitly reported or indirectly assessable in several of the included studies and was quantified using different metrics, including vector error, radial error, Euclidean error, trajectory deviation, and the spatial distance between the intended anatomical target and the final active stimulation contact.

One of the most comprehensive evaluations of stereotactic accuracy was reported by Burchiel et al. (2013), who assessed DBS electrode placement accuracy using MRI‑based targeting combined with intraoperative CT verification in a prospective cohort of 60 patients. Across 119 implanted electrodes, the mean vector error between the planned target and final electrode position was 1.59 ± 1.11 mm, while the mean deviation from the intended trajectory was 1.24 ± 0.87 mm. Interestingly, targeting accuracy differed across DBS targets, with Vim electrodes showing a mean vector error of approximately 1.9 mm, significantly greater than that observed for GPi implantation (1.29 mm; p = 0.01). The authors suggested that the relative proximity of the Vim target to the lateral ventricles may increase susceptibility to targeting deviation due to brain shift or trajectory constraints (Burchiel et al., 2013).

Comparable levels of stereotactic accuracy were reported by Chen et al. (2017) in a prospective cohort of 56 patients undergoing Vim DBS for essential tremor using indirect atlas‑based targeting without microelectrode recording. Electrode placement accuracy was quantified using both radial and Euclidean error measurements. The mean radial error was 0.9 ± 0.3 mm in the awake group and 0.9 ± 0.4 mm in the asleep group (p = 0.75), while Euclidean error measured 1.1 ± 0.6 mm and 1.2 ± 0.5 mm, respectively (p = 0.92). These findings demonstrated that sub‑millimetric to low‑millimetric targeting precision can be achieved using image‑guided stereotactic techniques even in the absence of intraoperative electrophysiological mapping (Chen et al., 2017).

Other studies evaluated the relationship between anatomical targeting and the functionally optimal stimulation site. In a series of 20 patients undergoing Vim DBS with direct anatomical targeting using a white‑matter attenuated inversion recovery (WAIR) sequence, Vassal et al. (2012) measured the spatial discrepancy between the inferior anatomical border of the Vim and the location of the clinically optimal stimulation contact. The mean distance between the anatomical Vim border and the effective stimulation site was 2.77 ± 2.1 mm (range 0–5.5 mm). Despite this anatomical‑functional discrepancy, tremor outcomes remained excellent, with approximately 90% improvement in tremor severity, suggesting that minor anatomical targeting deviations can be compensated during intraoperative stimulation testing.

Similarly, electrophysiological mapping studies have demonstrated that the functional boundaries of the Vim may not perfectly coincide with atlas‑based anatomical coordinates. Pedrosa et al. (2018) reported that functional microelectrode recordings identified tremor‑related neuronal activity across a depth range of approximately 5–12 mm along the planned trajectory, often requiring adjustments of the final electrode location based on intraoperative physiological responses.

Several imaging‑guided targeting studies also evaluated accuracy indirectly by comparing imaging‑defined targets with the final clinically effective stimulation site. For example, tractography‑based targeting approaches demonstrated that the centroid of the dentato‑rubro‑thalamic tract (DRTT) may lie several millimeters away from classical atlas‑based Vim coordinates. In a comparative tractography study, Yang et al. (2022) reported that the mean Euclidean distance between the final active contact and the probabilistic DRTT centroid was 3.32 ± 1.70 mm, compared with 5.01 ± 2.12 mm using deterministic tractography. Similarly, Sammartino et al. (2016) reported a mean targeting error of approximately 1.9 mm for DRTT‑based targeting compared with 2.8 mm for atlas‑based coordinates.

Other investigations have demonstrated that connectivity‑defined or patient‑specific targets may differ from traditional atlas coordinates by several millimeters. Ferreira et al. (2021) reported mean Euclidean distances of approximately 1.88–2.12 mm between connectivity‑defined Vim locations and atlas‑based targets across a large normative dataset, highlighting substantial interindividual anatomical variability.

Overall, across the included literature, reported targeting errors typically ranged between approximately 0.9 and 3 mm depending on the definition of accuracy and the targeting strategy used. These findings indicate that modern stereotactic techniques generally achieve sub‑millimetric to low‑millimetric precision, although the optimal functional stimulation site may not always coincide exactly with the anatomically defined Vim target.

**Imaging‑Based Targeting Approaches**

Considerable heterogeneity was observed across studies regarding imaging strategies used for Vim localization. Because the ventral intermediate nucleus cannot be reliably visualized on conventional MRI sequences, several indirect, structural, and connectivity‑based targeting approaches have been proposed.

The most widely used approach remains indirect atlas‑based targeting based on stereotactic coordinates relative to the AC–PC plane. Several studies in the present review used this method, typically referencing the Schaltenbrand–Wahren or Guiot coordinate systems (Papavassiliou et al., 2004; Chen et al., 2017; Gravbrot et al., 2020; Burchiel et al., 2013). In these approaches, the Vim target is commonly defined approximately 13–14 mm lateral to the midline and 25% of the AC–PC distance anterior to the posterior commissure, with adjustments based on ventricular width or thalamic morphology.

However, substantial anatomical variability of the Vim across individuals has been reported, raising concerns regarding the precision of purely atlas‑based targeting strategies. Structural connectivity studies have demonstrated that the functional motor thalamus may vary by several millimeters between individuals. Ferreira et al. (2021) showed that connectivity‑defined Vim locations differed from atlas‑derived coordinates by up to 5 mm in some subjects, highlighting the limitations of fixed stereotactic coordinates.

To address these limitations, several studies have investigated advanced imaging methods for direct or indirect visualization of the Vim. Vassal et al. (2012) demonstrated that a white‑matter attenuated inversion recovery (WAIR) MRI sequence at 1.5 T can directly visualize the Vim as a hypointense band within the ventrolateral thalamus, bordered posteriorly by the ventrocaudal nucleus and inferiorly by the prelemniscal radiations. This approach enabled direct anatomical targeting without reliance on atlas coordinates.

Ultra‑high‑field MRI techniques have also been explored to improve thalamic nucleus visualization. Najdenovska et al. (2019) reported that susceptibility‑weighted imaging at 7 T allows visualization of the Vim as a hyperintense structure within the ventrolateral thalamus and demonstrated improved spatial correspondence with histological atlases. Similarly, Su et al. (2020) showed that white‑matter‑nulled MPRAGE imaging at 7 T enables direct visualization and segmentation of the Vim, facilitating patient‑specific targeting for MR‑guided focused ultrasound thalamotomy.

Another major direction in imaging‑based targeting is the use of diffusion MRI tractography to identify the dentato‑rubro‑thalamic tract (DRTT), a key cerebello‑thalamo‑cortical pathway involved in tremor generation. Several studies have proposed targeting the thalamic portion of the DRTT rather than relying solely on atlas‑defined Vim coordinates (Sammartino et al., 2016; Fenoy and Schiess, 2018; Yang et al., 2022; Bruno et al., 2021). These approaches aim to account for patient‑specific anatomical variability by identifying the center of the DRTT within the thalamus.

Connectivity‑based thalamic segmentation methods represent another emerging strategy. Middlebrooks et al. (2018) demonstrated that probabilistic tractography can be used to parcellate the thalamus according to its cortical connectivity, allowing identification of thalamic regions connected to the primary motor cortex that correspond closely to the functional Vim. Similar approaches have been used to predict tremor improvement following DBS by analyzing the overlap between the volume of tissue activated and connectivity‑defined thalamic subregions.

Finally, automated multi‑atlas segmentation techniques have been developed to enable rapid identification of thalamic nuclei using structural MRI data. The THOMAS algorithm described by Su et al. (2019) demonstrated high segmentation accuracy for thalamic nuclei with Dice similarity coefficients of approximately 0.85 compared with expert manual segmentation.

Taken together, these studies illustrate the evolving landscape of imaging‑based targeting strategies for Vim interventions, ranging from traditional atlas‑based coordinates to advanced MRI visualization, tractography‑guided targeting, and connectivity‑based segmentation approaches.

**Risk of Bias Assessment**

The methodological quality of the included studies was evaluated using two established critical appraisal tools according to study design. The Joanna Briggs Institute (JBI) critical appraisal tools were applied to clinical observational studies, including case series, cohort studies, and analytical cross‑sectional designs, whereas the QUADAS‑2 tool was used for imaging and diagnostic‑accuracy–type studies. Overall, 25 studies were assessed for risk of bias, including 15 studies evaluated using JBI instruments and 10 studies assessed using QUADAS‑2.Overall risk‑of‑bias judgments indicated generally acceptable methodological quality across the included literature, although substantial variability was observed depending on study design and methodological focus. Across all studies, 11 were judged to have a low overall risk of bias, 10 were classified as having moderate risk, and 4 were considered to have high risk of bias. Studies assessed with the JBI tools generally demonstrated lower risk of bias compared with imaging‑methodological investigations assessed using QUADAS‑2. Most clinical observational studies evaluating surgical targeting or clinical outcomes following Vim interventions were rated as low risk of bias. These included Anthofer et al. (2014), Bardinet et al. (2010), Klostermann et al. (2003), Diaz et al. (2020), Middlebrooks et al. (2018), Vassal et al. (2012), Gravbrot et al. (2020), Burchiel et al. (2013), Bruno et al. (2021), and Pedrosa et al. (2018). These studies consistently reported clear patient selection criteria, well‑defined surgical or imaging procedures, and appropriate reporting of clinical or methodological outcomes. A moderate risk of bias was identified in several studies, primarily reflecting methodological limitations inherent to retrospective designs, incomplete reporting of outcome measurements, or heterogeneity in imaging analysis pipelines. Studies classified as moderate risk included Yang et al. (2015), Yang et al. (2022), Papavassiliou et al. (2004), Najdenovska et al. (2019), Tian et al. (2018), Su et al. (2020), Fenoy and Schiess (2018), Chen et al. (2018), Goedemans et al. (2024), and Sammartino et al. (2016). In these studies, potential bias frequently arose from incomplete reporting of participant selection procedures, limited description of reference standards in imaging validation, or variability in follow‑up duration and outcome assessment. Four studies were considered to have a high risk of bias, largely among imaging‑focused methodological investigations. These included Young et al. (2009), Su et al. (2019), Hyam et al. (2012), and Ferreira et al. (2021). The principal contributors to high risk ratings were related to patient selection bias, lack of clearly defined reference standards for imaging validation, and incomplete reporting of study flow and timing between index tests and reference assessments. Domain‑level assessment of the QUADAS‑2 studies revealed that the most frequent concerns arose in the patient‑selection and reference‑standard domains. Several imaging studies relied on small convenience samples or retrospective imaging datasets, which increased the potential for selection bias. Additionally, in a number of methodological investigations, the reference standard for defining Vim boundaries or validating segmentation algorithms was not clearly defined or relied on indirect anatomical assumptions rather than histological confirmation. Interpretation of the risk‑of‑bias heatmaps further illustrates these patterns. The JBI heatmap demonstrates that most clinical studies achieved high scores in the “Yes” domain, indicating that the majority of appraisal criteria were satisfied. Many studies scored between 8 and 10 positive responses across the checklist items, supporting a low overall risk classification. Only a small number of JBI‑assessed studies showed notable numbers of “Unclear” responses, typically reflecting insufficient methodological reporting rather than clear methodological flaws. In contrast, the QUADAS‑2 heatmap demonstrates greater heterogeneity across methodological domains. Several imaging‑methodological studies showed high or unclear risk in the patient selection and reference standard domains, reflecting uncertainties regarding dataset representativeness and validation of imaging‑derived targets. However, many studies demonstrated low risk in the flow and timing domain, suggesting that the imaging analysis and outcome measurements were generally performed within consistent methodological frameworks. Taken together, the overall risk‑of‑bias assessment suggests that while most clinical observational studies evaluating Vim targeting and tremor surgery exhibit acceptable methodological quality, imaging‑methodological studies introduce greater heterogeneity and potential bias related to dataset selection and validation strategies. These methodological differences should be considered when interpreting the collective evidence regarding imaging‑based targeting approaches for the ventral intermediate nucleus.

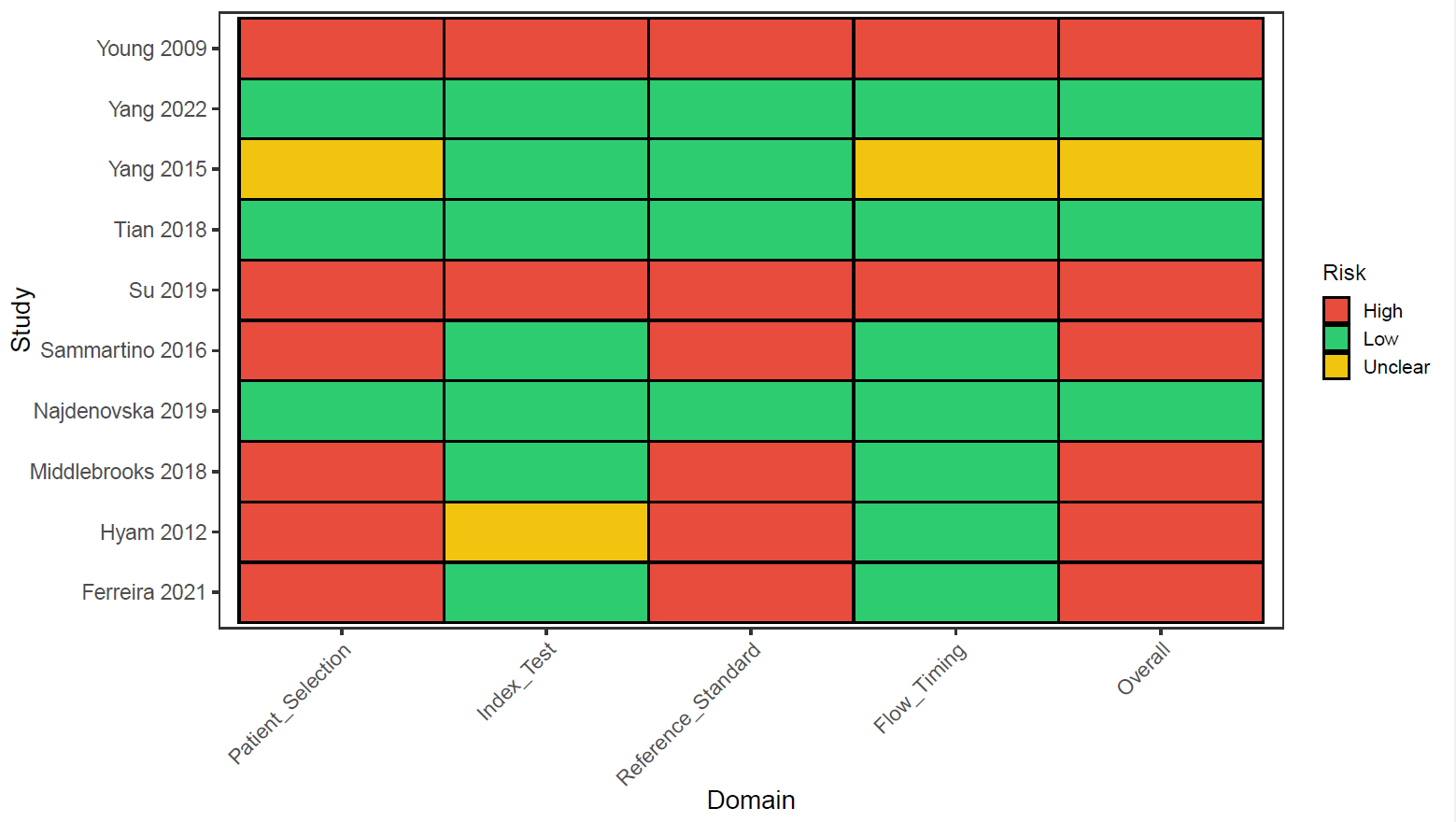

Figure 2: risk of bias assessment by QURADS-2 checklist.

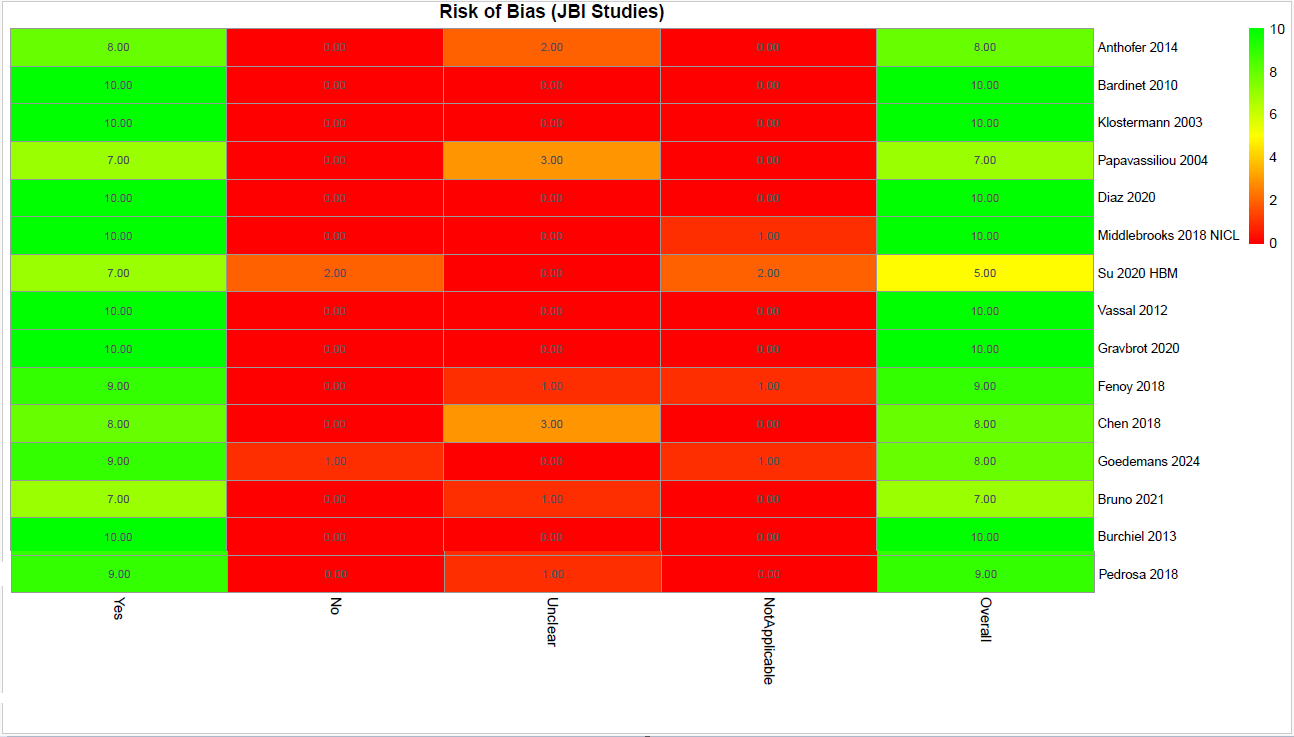

Figure3: risk of bias assessment by JBI checklist.

**Eligibility for Quantitative Synthesis**

Among 25 included studies for this systematic review, 7 provided sufficient quantitative outcome data from 6 studies to be included in the meta‑analysis of tremor improvement. Eligibility for quantitative synthesis required studies to report extractable clinical outcome data, including preoperative and postoperative tremor severity scores measured using validated rating scales, together with sufficient statistical information to estimate measures of central tendency and variability. Specifically, studies were required to report either mean ± standard deviation values or convertible summary statistics (e.g., median and range) for tremor severity before and after intervention. The majority of the included studies were not eligible for meta‑analysis because their primary objectives focused on surgical technique, targeting accuracy, electrophysiological characterization, or imaging‑based methodological investigations rather than clinical tremor outcomes. Consequently, these studies did not report quantitative tremor rating scale data required for effect size calculation. For example, Burchiel et al. (2013) evaluated the stereotactic accuracy of deep brain stimulation electrode placement using intraoperative CT verification without microelectrode recording in a cohort of 60 patients. The study reported detailed surgical accuracy metrics, including vector error (mean 1.59 ± 1.11 mm) and trajectory deviation, but did not report preoperative or postoperative tremor severity scores. As a result, the study was not eligible for inclusion in the quantitative meta‑analysis. Similarly, Pedrosa et al. (2018) conducted an intraoperative electrophysiological mapping study of tremor‑related neuronal activity in the ventrolateral thalamus during DBS implantation in patients with essential tremor. Although the study provided detailed neurophysiological data derived from microelectrode recordings, it did not report clinical outcome measures or tremor rating scale scores and therefore could not contribute to quantitative outcome synthesis. In other cases, studies reported clinical improvement only as percentage change without providing baseline and postoperative scores or measures of variability. For instance, Diaz et al. (2020) reported a mean tremor improvement of 79.7% ± 22.4% following individualized anatomy‑based VIM‑cZI targeting in patients with essential tremor. However, because baseline and follow‑up tremor severity scores were not reported, it was not possible to compute standardized effect sizes for inclusion in the meta‑analysis. Additionally, several studies investigated alternative treatment modalities rather than deep brain stimulation. For example, Su et al. (2020) evaluated patient‑specific imaging‑based targeting for ventral intermediate nucleus ablation using magnetic resonance–guided focused ultrasound (MRgFUS). Although the study examined imaging predictors of clinical outcome, it did not report extractable pre‑ and postoperative tremor rating scores and involved a different therapeutic modality, rendering it ineligible for inclusion in the DBS outcome meta‑analysis. Among the studies included in the systematic review, only four investigations reported sufficiently detailed tremor outcome data to permit quantitative synthesis. These studies provided extractable pre‑ and postoperative tremor severity scores measured using validated clinical scales, allowing calculation of standardized mean differences for pre‑ to post‑treatment comparisons. The limited number of studies eligible for quantitative synthesis reflects the substantial methodological heterogeneity within the literature on Vim targeting. Many investigations prioritize anatomical targeting strategies, imaging validation, or intraoperative physiology rather than standardized clinical outcome reporting, which restricts their inclusion in meta‑analytic outcome assessment.

**Pre–Post Tremor Improvement (Primary Meta-analysis)**

Across the six studies included in the quantitative synthesis, comprising seven independent cohorts, tremor severity demonstrated a consistent and substantial improvement from baseline to postoperative assessment. Despite heterogeneity in study design, tremor rating scales, and anatomical targeting strategies within the ventral intermediate nucleus (Vim) region, all cohorts reported reductions in tremor severity following intervention. Standardized mean change scores were calculated from pre–post comparisons to quantify treatment effects within each cohort. Across all included datasets, standardized mean differences (SMDs) were uniformly large and negative, reflecting pronounced postoperative tremor reduction. Notably, the magnitude of improvement remained relatively consistent across studies despite the use of different clinical assessment scales, including Fahn–Tolosa–Marín Tremor Rating Scale (FTMTRS/CRST) and TETRAS-based measures. Using a random‑effects model to account for between‑study variability, the pooled analysis demonstrated overall treatment effect (SMD −3.91, 95% CI −4.81 to −3.01; p < 0.0001). Importantly, the confidence intervals for all individual cohorts and the pooled estimate remained entirely below zero, confirming statistically significant improvement in tremor severity across the included literature. (Figure 4)

**Heterogeneity Analysis**

Between‑study heterogeneity was evaluated using Cochran’s Q statistic and the I² metric. The analysis revealed statistically significant heterogeneity (Q = 18.12, p = 0.0059), corresponding to an I² value of 66.9% and a between‑study variance (τ²) of 0.63. These findings indicate that approximately two‑thirds of the observed variability in effect sizes reflects true differences between studies rather than random sampling error. Several factors likely contribute to this heterogeneity, including differences in patient populations, tremor etiology, baseline tremor severity, surgical targeting approaches within the Vim complex, and variability in postoperative follow‑up intervals. Nonetheless, despite these methodological and clinical differences, the direction of treatment effect remained consistent across all cohorts. To further characterize the expected variability in clinical outcomes, the random‑effects model yielded a wide but consistently negative prediction interval, suggesting that future studies conducted under similar conditions would also be expected to demonstrate substantial tremor reduction following intervention.

**Sensitivity Analyses**

strength of the pooled estimate was evaluated using leave‑one‑out sensitivity analyses. Sequential exclusion of each cohort resulted in pooled standardized mean differences ranging from −3.73 to −4.19, with all estimates remaining statistically significant. Importantly, no single study materially altered the magnitude, direction, or statistical significance of the pooled treatment effect. These findings indicate that the observed improvement in tremor severity was stable and not disproportionately driven by any individual dataset. (Figure 5)

**Assessment of Small‑Study Effects**

Potential small‑study effects were explored through visual inspection of funnel plots constructed using standardized mean differences and corresponding standard errors. The distribution of effect estimates appeared broadly symmetrical around the pooled effect size, without clear evidence of asymmetry suggestive of publication bias. (Figure 8) Trim‑and‑fill analysis did not identify any potentially missing studies (k₀ = 0), and the pooled effect estimate remained unchanged following adjustment. However, because the number of included studies remained below the commonly recommended threshold for reliable asymmetry testing, these findings should be interpreted cautiously. (Figure 9)

**Influence Diagnostics**

Influence diagnostics were conducted to determine whether any individual cohort exerted disproportionate influence on the pooled estimate or the observed heterogeneity. Leave‑one‑out analyses confirmed that removal of individual studies did not substantially alter the pooled effect size. (Figure7)

Complementary evaluation using Baujat plots demonstrated that while certain cohorts contributed modestly to overall heterogeneity, no single study simultaneously exhibited high influence on the pooled estimate and high contribution to heterogeneity. (Figure6) Additional influence diagnostics, including standardized residuals, Cook’s distance, and DFFITS values, similarly indicated that none of the included cohorts exceeded conventional thresholds for undue influence.

Taken together, these analyses support the robustness and internal consistency of the meta‑analytic findings, reinforcing the conclusion that tremor severity improves markedly following intervention across diverse study settings and targeting methodologies.

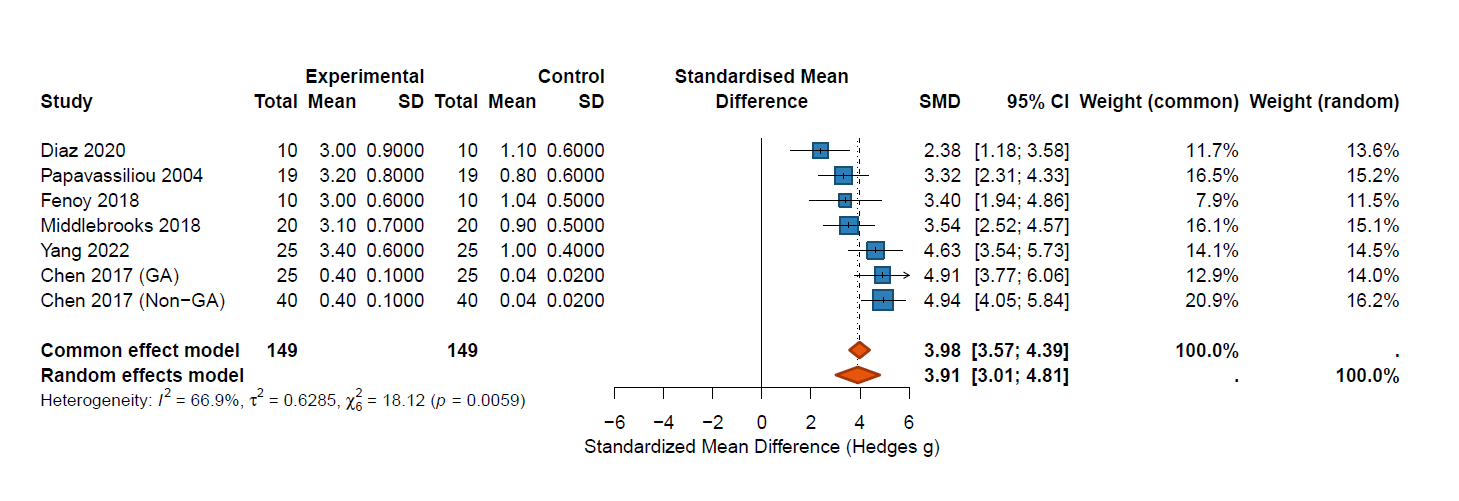

Figure 4. Forest plot of the meta‑analysis demonstrating the pooled standardized mean difference in tremor scores between baseline and post‑treatment measurements. Each square represents the effect size for an individual study, with the size of the square proportional to study weight. Horizontal lines indicate 95% confidence intervals. The diamond represents the pooled effect estimate from the random‑effects model.

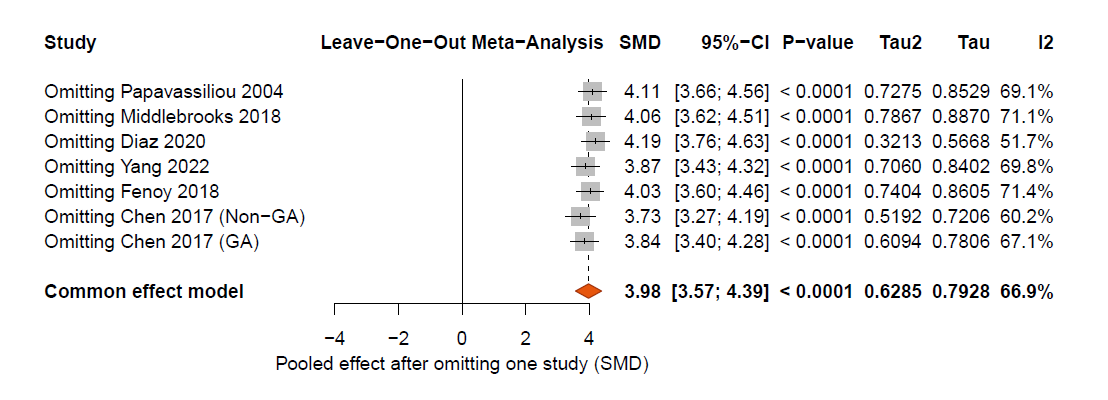

Figure 5. Leave‑one‑out sensitivity analysis showing pooled standardized mean differences after sequential removal of each study. The stability of the pooled effect size across all iterations indicates that the overall result was not driven by any single study.

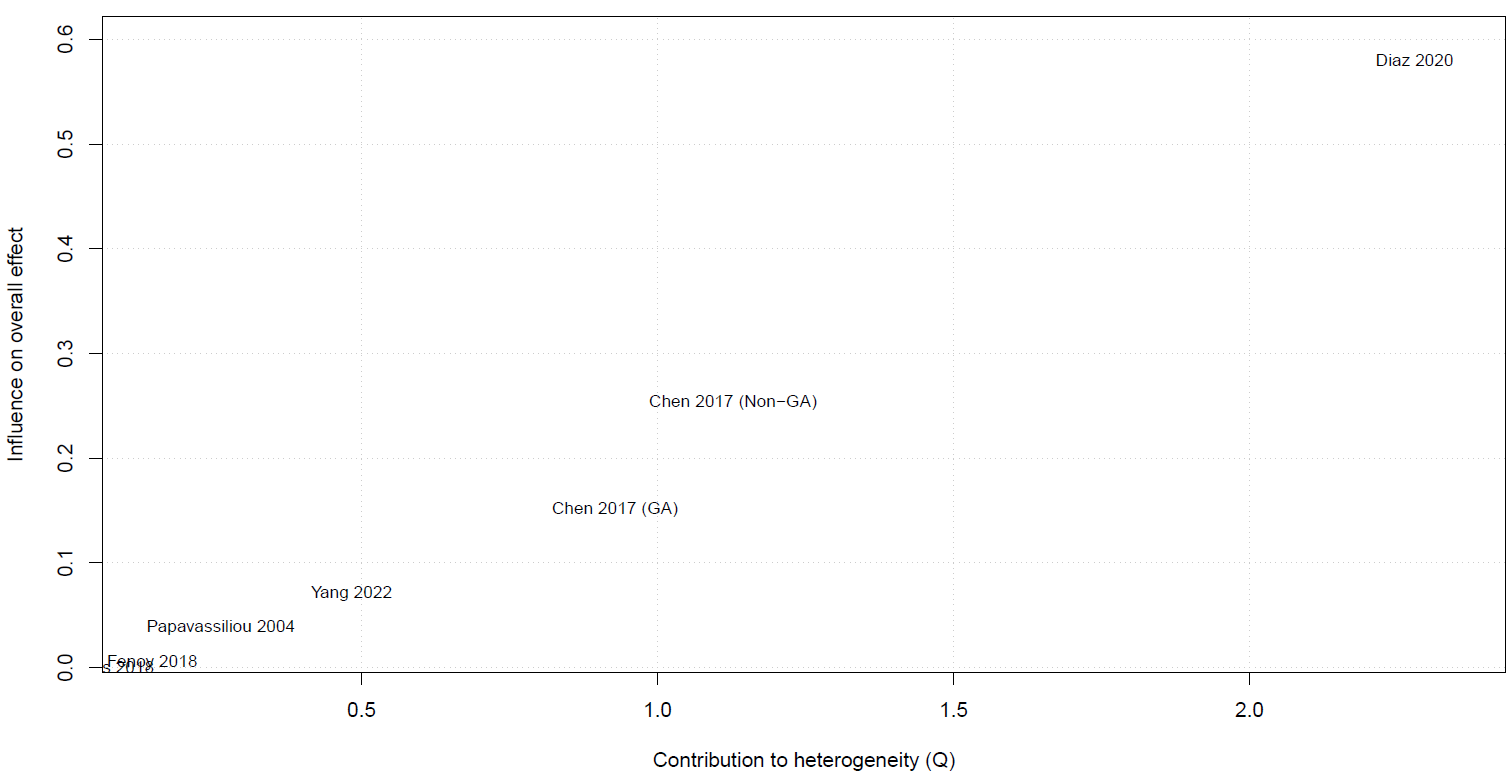

Figure 6. Baujat plot illustrating the contribution of each study to overall heterogeneity and influence on the pooled effect size. The study by Diaz et al. (2020) showed the largest contribution to heterogeneity but did not represent a statistical outlier.

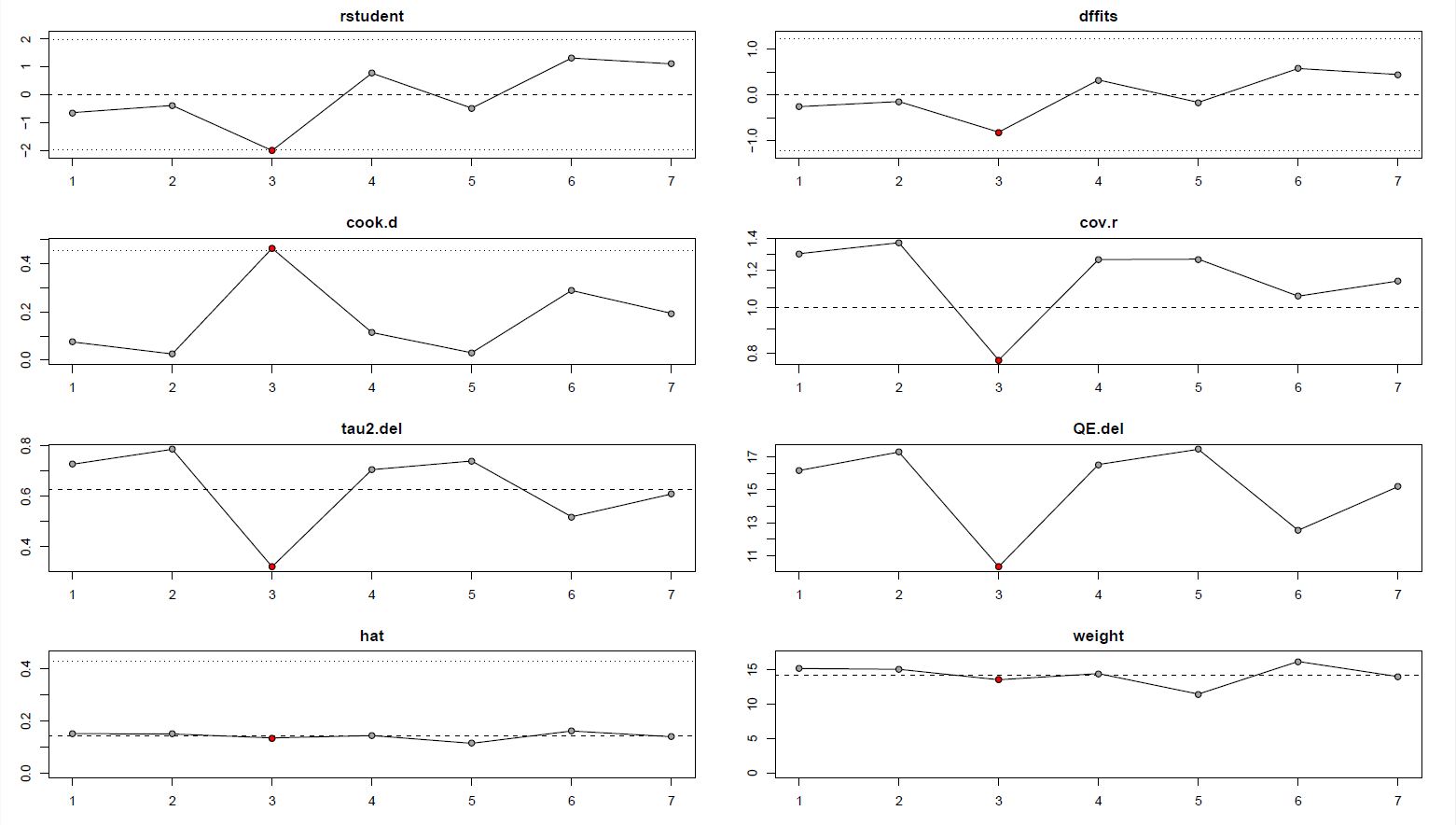

Figure 7. Influence diagnostics evaluating the impact of individual studies on the meta‑analysis model. Metrics include studentized residuals, Cook’s distance, DFFITS, covariance ratios, hat values, and changes in heterogeneity statistics after study deletion.
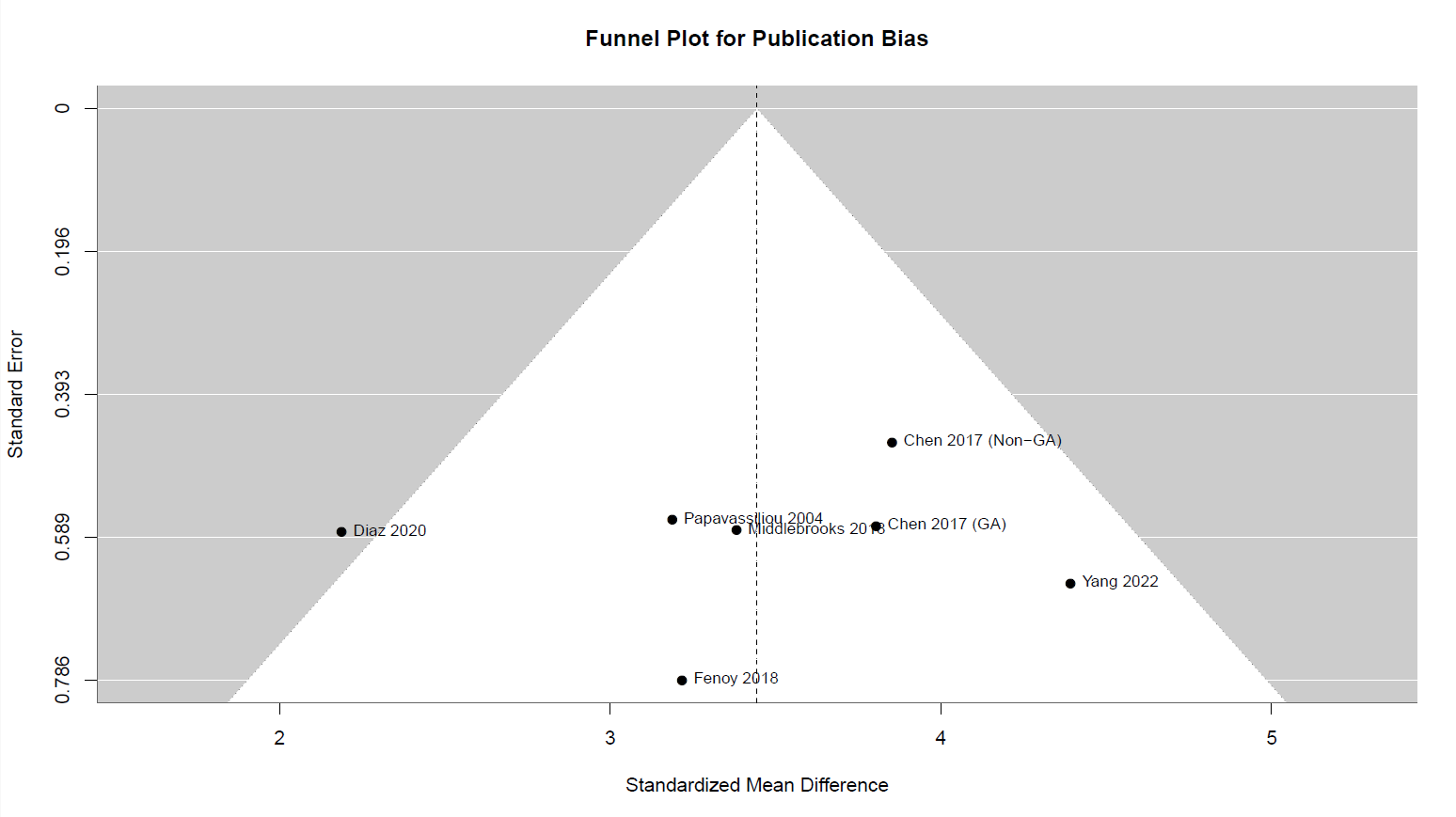

Figure 8. Funnel plot assessing potential publication bias in the included studies. The distribution of studies appears broadly symmetric around the pooled effect estimate.

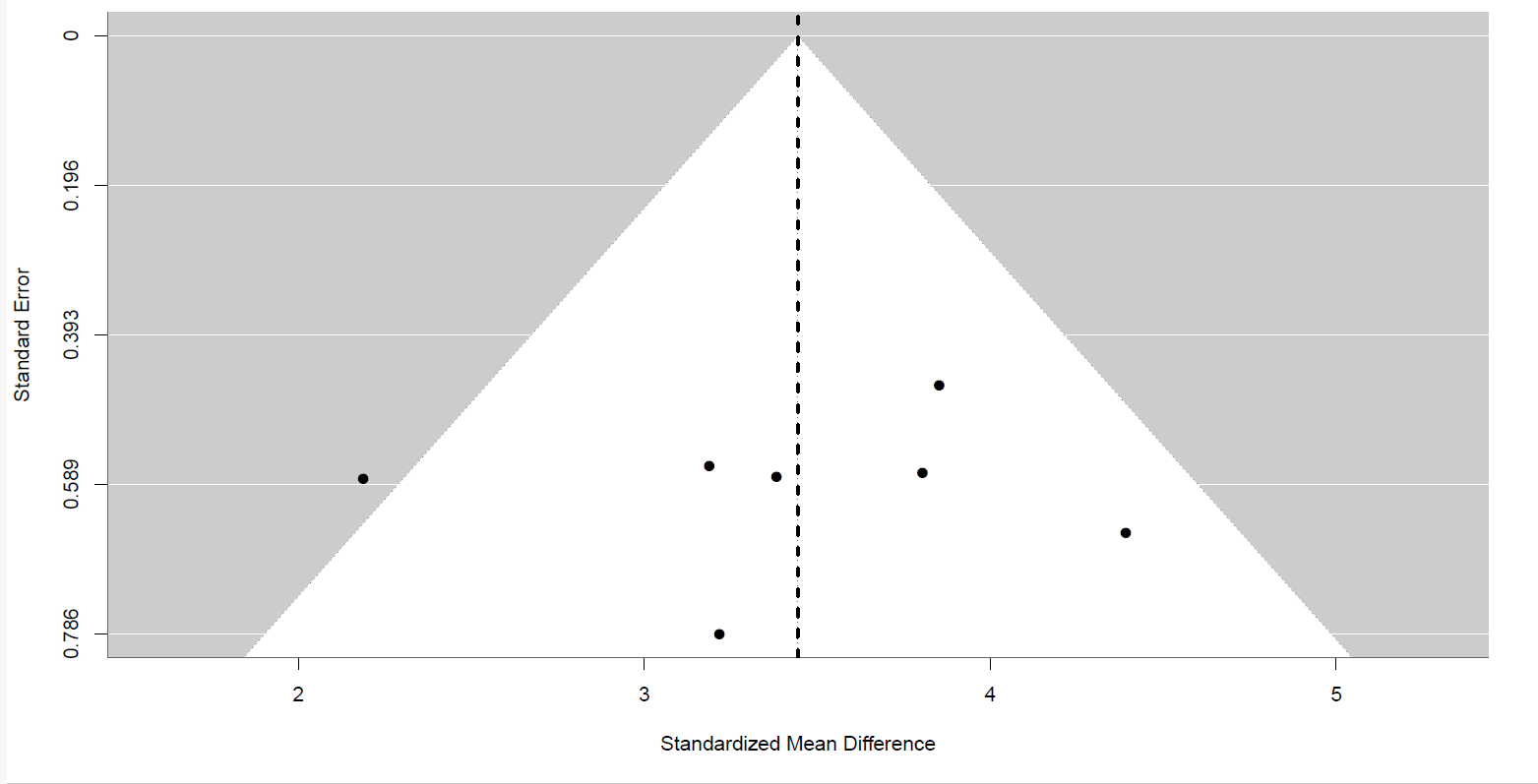

Figure 9. Trim and Fill plot., although with considering study number limitation, this plot did not identify any potentially missing studies (k₀ = 0).
