## Supplementary material for "Tremor Improvement Despite Heterogeneous Ventral Intermediate Nucleus Targeting in Deep Brain Stimulation: A Systematic Review and Meta-Analysis": protocol prospero

NIHR | National Institute for Health and Care Research

PROSPERO
International prospective register of systematic reviews

### Tremor Improvement Despite Heterogeneous Ventral Intermediate Nucleus Targeting in Deep Brain Stimulation: A Systematic Review and Meta Analysis

*Farzan Fahim, amirmahdi mojtahedzadeh*

##### Citation

Farzan Fahim, amirmahdi mojtahedzadeh. Tremor Improvement Despite Heterogeneous Ventral Intermediate Nucleus Targeting in Deep Brain Stimulation: A Systematic Review and Meta Analysis. PROSPERO 2026 CRD420261362914. Available from https://www.crd.york.ac.uk/PROSPERO/view/CRD420261362914.

#### REVIEW TITLE AND BASIC DETAILS

##### Review title

Tremor Improvement Despite Heterogeneous Ventral Intermediate Nucleus Targeting in Deep Brain Stimulation: A Systematic Review and Meta Analysis

##### Condition or domain being studied

*Deep Brain Stimulation; Stereotactic Operation On Thalamus; Tremor*

##### Rationale for the review

Deep brain stimulation (DBS) targeting the ventral intermediate nucleus (Vim) of the thalamus is a well-established treatment for medically refractory tremor, particularly in essential tremor and Parkinson's disease. However, the Vim cannot be reliably visualized on conventional MRI, leading to the development of multiple targeting strategies, including atlas-based stereotactic coordinates, advanced MRI visualization, diffusion tractography of the dentato-rubro-thalamic tract, and connectivity-based thalamic segmentation. Despite these advances, no consensus exists regarding the optimal targeting method or its relationship to clinical outcomes. This systematic review and meta-analysis aims to synthesize current evidence on Vim targeting strategies and evaluate their association with postoperative tremor improvement.

##### Review objectives

The primary objective of this systematic review and meta-analysis is to evaluate and synthesize the existing evidence on strategies used to localize and target the ventral intermediate nucleus (Vim) of the thalamus in patients undergoing deep brain stimulation (DBS) for tremor disorders. Specifically, the review aims to compare anatomical, imaging-based, tractography-guided, connectivity-based, and electrophysiological targeting approaches and assess their relationship with clinical outcomes. A secondary objective is to quantitatively evaluate pre- to postoperative tremor improvement across studies reporting standardized tremor scores and to determine whether different targeting methodologies are associated with consistent therapeutic effects despite substantial methodological heterogeneity.

##### Keywords
Deep brain stimulation; Tremor; Systematic review

##### Country
Iran (Islamic Republic of)

#### ELIGIBILITY CRITERIA

##### Population

*Included*
patients who undergo dbs to target vim

##### Intervention(s) or exposure(s)

*Included*
Ventral Intermediate Nucleus target

##### Comparator(s) or control(s)
This review does not have any comparators

##### Study design
Only nonrandomized study types will be included.

*Included*
cohorts

cross sectional

case control

##### Context
The review includes studies evaluating localization or targeting of the ventral intermediate nucleus (Vim) of the thalamus in patients undergoing surgical treatment for tremor, primarily deep brain stimulation. Eligible populations consist mainly of adults with medically refractory tremor disorders, most commonly essential tremor and Parkinson's disease tremor. Studies were conducted in clinical neurosurgical settings as well as imaging methodological environments. Targeting approaches include atlas-based stereotactic coordinates, microelectrode recording–guided physiological mapping, advanced MRI visualization (e.g., WAIR or 7T imaging), diffusion MRI tractography of the dentato-rubro-thalamic tract, and connectivity-based thalamic segmentation.

Observational cohorts, prospective studies, cross-sectional studies, and large case series (≥10 patients) were eligible.

#### TIMELINE OF THE REVIEW

##### Date of first submission to PROSPERO
07 April 2026

##### Review timeline
Start date: 1 September 2025. End date: 1 June 2026.

##### Date of registration in PROSPERO
07 April 2026

#### AVAILABILITY OF FULL PROTOCOL

##### Availability of full protocol
A full protocol has not been written.

#### SEARCHING AND SCREENING

##### Search for unpublished studies
Only published studies will be sought.

##### Main bibliographic databases that will be searched
The main databases to be searched are *Embase.com*, *PubMed* and *Scopus*.

##### Search language restrictions
There are no language restrictions.

##### Search date restrictions
There are no search date restrictions.

##### Other methods of identifying studies
No other methods will be used.

##### Link to search strategy
A full search strategy has been uploaded to PROSPERO. The PDF may be accessed through this link https://www.crd.york.ac.uk/PROSPEROFILES/c6efd30b6069759712283517ecdbfbbf.pdf.

##### Selection process
Studies will be screened independently by at least two people (or person/machine combination) with a process to resolve differences.

##### Other relevant information about searching and screening
None

#### DATA COLLECTION PROCESS

##### Data extraction from published articles and reports

Data will be extracted independently by at least two people (or person/machine combination) with a process to resolve differences.

Authors will not be contacted for further information.

##### Study risk of bias or quality assessment

Risk of bias will be assessed using: *QUADAS-2*

Data will be assessed independently by at least two people (or person/machine combination) with a process to resolve differences.

Additional information will be sought from study investigators if required information is unclear or unavailable in the study publications/reports.

##### Reporting bias assessment

Risk of bias due to missing results will be assessed

##### Certainty assessment

Certainty of findings will not be assessed

#### OUTCOMES TO BE ANALYSED

##### Main outcomes

tremor improvement is the main outcome

##### Additional outcomes

vim targeting accuracy

image base targeting accuravy

#### PLANNED DATA SYNTHESIS

##### Strategy for data synthesis

tremor improvement from primary studies will extracted and quantitively synthesis will run, if scales of measuring of tremor improvement are differents, we wil, use smd with ci 95% to evaluate our main outcome

#### CURRENT REVIEW STAGE

##### Stage of the review at this submission

| Review stage | Started | Completed |
|---|---|---|
| Pilot work | ✓ | ✓ |

| Review stage | Started | Completed |
|---|---|---|
| Formal searching/study identification | ✓ | |
| Screening search results against inclusion criteria | ✓ | |
| Data extraction or receipt of IPD | | |
| Risk of bias/quality assessment | | |
| Data synthesis | | |

##### Review status
The review is currently planned or ongoing.

##### Publication of review results
Results of the review will be published.

#### REVIEW AFFILIATION, FUNDING AND PEER REVIEW

##### Review team members
**Dr Farzan Fahim** (review guarantor and contact) ORCID: 0000-0003-0591-674X. Shohada-E-Tajrish Hospital. Iran.

No conflict of interest declared.

**Dr amirmahdi mojtahedzadeh**. shohada-e-tajrish hospital. Iran.

No conflict of interest declared.

##### Named contact

##### Review affiliation
shahid beheshti university of medical science

##### Funding source
Review has no specific/external funding but is supported by guarantor/review team (non-commercial) institutions.

##### Peer review
There has been no peer review of this planned review.

#### ADDITIONAL INFORMATION

##### Review conflict of interest
Declared individual interests are recorded under team member details.. No additional interests are recorded for this review.

##### Medical Subject Headings
Deep Brain Stimulation; Tremor; Thalamic Nuclei

#### SIMILAR REVIEWS

##### Check for similar records already in PROSPERO

there is no similar articles in the specific field that we have choosen

##### PROSPERO version history

- Version 1.0, published 07 Apr 2026

##### Disclaimer

The content of this record displays the information provided by the review team. PROSPERO does not peer review registration records or endorse their content.

PROSPERO accepts and posts the information provided in good faith; responsibility for record content rests with the review team. The guarantor for this record has affirmed that the information provided is truthful and that they understand that deliberate provision of inaccurate information may be construed as scientific misconduct.

PROSPERO does not accept any liability for the content provided in this record or for its use. Readers use the information provided in this record at their own risk.

Any enquiries about the record should be referred to the named review contact
