## Supplementary material for "Tremor Improvement Despite Heterogeneous Ventral Intermediate Nucleus Targeting in Deep Brain Stimulation: A Systematic Review and Meta-Analysis": search strategy

Serach Strategy

**Pubmed**

("ventral intermediatenucleus"[tiab] OR VIM[Title/Abstract] OR "ventralisintermedius"[ tiab] OR "thalamic nucleus"[ tiab]) AND("segmentation" OR "parcellation" OR "targeting"OR "localization" OR "atlas" OR "tractography" OR"diffusion MRI" OR "connectivity" OR "automatedsegmentation" OR "artificial intelligence"OR"Electrophysiologic mapping" OR "brain mapping" OR mappingOR parcel* OR segment* OR tractograph* OR diffus* OR connect* OR fMRI OR FGATIROR "fast gray matter acquisition T1 inversion recovery" ) AND("deep brain stimulation"[tiab] OR "deep brainstimulation"[ tiab] OR DBS[tiab])

**Scopus:**

(TITLE-ABS-KEY("ventral intermediatenucleus" OR VIM OR "ventralis intermedius" OR "thalamicnucleus")) AND(TITLE-ABS-KEY("segmentation" OR"parcellation" OR "targeting" OR "localization"OR "atlas" OR "tractography" OR "diffusion MRI"OR "connectivity" OR "automated segmentation" OR"artificial intelligence" OR "Electrophysiologic mapping"OR "brain mapping" OR mapping OR parcel* OR segment* OR tractograph*OR diffus* OR connect* OR fMRI OR FGATIR OR "fast gray matter acquisitionT1 inversion recovery")) AND (TITLE -AB S-KEY("deep brainstimulation" OR DBS))

**WOS:**

TS=("ventral intermediate nucleus"OR VIM OR "ventralis intermedius" OR "thalamic  nucleus") AND TS=("segmentation" OR"parcellation" OR "targeting" OR "localization"OR "atlas" OR "tractography" OR "diffusion MRI"OR "connectivity" OR "automated segmentation" OR"artificial intelligence" OR "Electrophysiologic mapping"OR "brain mapping" OR mapping OR parcel* OR segment* OR tractograph*OR diffus* OR connect* OR fMRI OR FGATIR OR "fast gray matter acquisitionT1 inversion recovery") AND TS=("deep brain stimulation" ORDBS)

**Embase:**

('ventral intermediate nucleus':ti,ab OR vim:ti,ab OR 'ventralis intermedius':ti,ab OR 'thalamic nucleus':ti,ab) AND ('segmentation':ti,ab OR 'parcellation':ti,ab OR 'targeting':ti,ab OR 'localization':ti,ab OR 'atlas':ti,ab OR 'tractography':ti,ab OR 'diffusion mri':ti,ab OR 'connectivity':ti,ab OR 'automated segmentation':ti,ab OR 'artificial intelligence':ti,ab OR 'electrophysiologic mapping':ti,ab OR 'brain mapping':ti,ab OR mapping:ti,ab OR parcel*:ti,ab OR segment*:ti,ab OR tractograph*:ti,ab OR diffus*:ti,ab OR connect*:ti,ab OR fmri:ti,ab OR fgatir:ti,ab OR 'fast gray matter acquisition t1 inversion recovery':ti,ab) AND ('deep brain stimulation':ti,ab OR dbs:ti,ab)
